# Mental health status and related factors influencing healthcare workers during the COVID-19 pandemic: a systematic review and meta-analysis

**DOI:** 10.1101/2023.07.21.23292948

**Authors:** Jia Huang, Zhu-Tang Huang, Xin-Ce Sun, Ting-Ting Chen, Xiao-Tian Wu

## Abstract

**Background:** The mental health of healthcare workers during the coronavirus-2019 pandemic was seriously affected, and the risk of mental health problems was high. The present study sought to systematically evaluate the mental health problems of healthcare workers worldwide during the pandemic and to determine the latest global frequency of COVID-19 associated mental health problems.

**Methods:** Data in the Cumulative Index to Nursing and Allied Health Literature (CINAHL), EMBASE, Elsevier, MEDLINE, PubMed, PsycINFO and the Web of Science before November 11, 2022, were systematically searched. Cohort, case-control and cross-sectional studies were included. The meta-analysis used a random effects model to synthesize the comprehensive prevalence rate of mental health problems. Subgroup analyses were performed based on time of data collection; whether the country was or was not developed; continent; doctors and nurses; doctors/nurses vs. other healthcare workers; and psychological evaluation scale.

**Results:** A total of 161 studies were included, including 341,014 healthcare workers worldwide, with women accounting for 82.8%. Occupationally, 16.2% of the healthcare workers were doctors, 63.6% were nurses and 13.3% were other medical staff. During the pandemic, 47% (95% confidence interval [CI], 35-60%) of healthcare workers reported job burnout, 38% (95% CI, 35-41%) experienced anxiety, 34% (95% CI 30-38%) reported depression, 30% (95% CI, 29-31%) had acute stress disorder, and 26% (95% CI, 21-31%) had post-traumatic stress disorder.

**Conclusions:** The study found that there were common mental health problems among health care workers during the COVID-19 pandemic. The most common was job burnout, followed by anxiety, depression, acute stress and post-traumatic stress disorder. Although the global pandemic has been brought under control, its long-term impact on the mental health of healthcare workers cannot be ignored. Additional research is required to develop measures to prevent, monitor and treat psychological disorders among healthcare workers.

## 1. Introduction

On January 9, 2020, a kind of novel coronavirus 2019-NCoV was officially identified as the pathogen of a viral pneumonia outbreak in Wuhan, China [1]. As of January 19, 2023, the cumulative number of confirmed cases of COVID-19 worldwide has reached 663,001,898, including 6,707,959 deaths, and these numbers are still increasing [2].

The pandemic situation of COVID-19, like other pandemics, has had a significant impact on the mental health of ordinary people, but because of the particularity of the working conditions of medical staff, they suffered a greater impact on their mental health [3]. Lack of treatment experience and protective equipment, understaffing and work overload, and ethical dilemmas in allocating scarce resources to other patients who also needed help had deleterious effects on the mental health of healthcare workers. Other factors, such as the potential risk of infecting family members and the and the emergency deployment of many non-respiratory and infectious disease professionals to front-line intensive care units with high risk in the face of a sharp increase in the number of patients, not only put healthcare workers worldwide at higher risk of infection, but caused great psychological stress [4–7]. Studies of the SARS pandemic and MERS epidemic showed that these psychological stresses increased the risk of mental health diseases among healthcare workers, including anxiety, depression, post-traumatic stress symptoms and job burnout [8,9]. These conditions not only had adverse effects on healthcare workers, but had serious adverse consequences for their co-workers, families, and society in general [10].

Many articles on this subject have been published recently, most of which showed an increase in the prevalence of mental diseases among health workers, but most of these studies were either aimed at all social people or people in non-medical fields or in specific medical personnel, such as front-line personnel, psychiatric professionals, rehabilitation areas, and some studies were carried out in specific countries and regions [11–15]. At present, the literature search deadline of the published research is mostly in 2020 or 2021 [16–19], and there were few studies included health care worker prevalence published in 2022 [20]. Because of the ongoing nature of the COVID-19 pandemic and the continuous change of the global pandemic situation, the mental health status of global healthcare workers is in a state of dynamic change. Our analysis, which included studies published through November 2022, evaluated the existing scientific evidence on the mental health status of medical staff during the COVID-19 pandemic. These findings can therefore provide the latest estimates on the rates of depression, anxiety, burnout, acute stress and post-traumatic stress disorder among healthcare workers.

This study conducted an innovative subgroup analysis according to the time of data collection included in the article. Compared with the previous subgroup analysis based on the publication time of the included articles, our findings can more accurately determine the mental health status of health care workers around the world at different times.

## 2. Materials and Methods

This systematic review and meta-analysis adhered to the Preferred Reporting Items for Systematic Reviews and Meta-Analyses (PRISMA) guidelines [21], and detailed information is provided in **S1 Table**. The study protocol has been registered in the International Prospective Register of Systematic Reviews (PROSPERO) as No. CRD42023392547.

### 2.1. Search strategy and selection criteria

To find all relevant studies, two independent researchers (J. Huang and T. T. Chen) searched the Cumulative Index to Nursing and Allied Health Literature (CINAHL), EMBASE, Elsevier, MEDLINE, PubMed, PsycINFO and Web of Science databases for all studies published through November 11, 2022, using medical subject word (MeSH) terms and specific keywords. The specific keywords used to search the databases included COVID-19 OR COVID 19 OR COVID 19 OR Infection SARS-CoV-2 OR SARS CoV 2 Infection OR SARS-CoV-2 Infections OR 2019 Novel Coronavirus Disease OR 2019 Novel Coronavirus Infection OR 2019-nCoV Disease OR 2019 nCoV Disease OR 2019-nCoV Diseases OR Disease, 2019-nCoV OR COVID-19 Virus Infection OR COVID-19 Virus Infection OR COVID-19 Virus Infections OR Infection, COVID-19 Virus OR Virus Infection, COVID-19 OR Coronavirus Disease 2019 OR Disease 2019, Coronavirus OR Coronavirus Disease-19 OR Coronavirus Disease 19 OR Severe Acute Respiratory Syndrome Coronavirus 2 Infection OR SARS Coronavirus 2 Infection OR COVID-19 Virus Disease OR COVID 19 Virus Disease OR COVID-19 Virus Diseases OR Disease, COVID-19 Virus OR Virus Disease, COVID-19 OR 2019-nCoV Infection OR 2019 nCoV Infection OR 2019-nCoV Infections OR Infection, 2019-nCoV OR COVID19 OR COVID-19 Pandemic OR COVID 19 Pandemic OR Pandemic, COVID-19 OR COVID-19 Pandemics AND Mental Health OR Health, Mental OR Mental Hygiene OR Hygiene, Mental AND Personnel, Health OR Healthcare Providers OR Healthcare Provider OR Provider, Healthcare OR Healthcare Workers OR Healthcare Worker OR Healthcare Professionals OR Healthcare Professional OR Professional, Healthcare OR “Health Personnel”. The retrieval strategy is described in detail in the supplementary document (**Retrieval strategy**).

### 2.2. Eligibility criteria

All the retrieved articles were screened independently by two researchers (J. Huang and T.T. Chen). Title and abstract were screened initially, with all articles that did not meet the inclusion criteria excluded. The full texts of the remaining articles were screened subsequently.

Criteria for inclusion in the systematic review and meta-analysis were: (a) studies of professionals who worked as healthcare workers during the COVID-19 pandemic; (b) cohort, cross-sectional and case-control studies; (c) studies that used a validated structured assessment scale; (d) studies of sample size ≥ 100 persons; and (e) studies with original and independent data. In addition, studies that reported mental health outcomes during the pandemic had to include at least one of the following categories: anxiety, depression, job burnout, post-traumatic stress disorder and acute stress disorder, with the prevalence rate being reported or able to be calculated. Studies were excluded if they reporting results from people other than healthcare workers, such as the general public, students, or police or were written in a language other than English.

### 2.3. Data extraction

Two researchers (X.C. Sun and Z.T. Huang) comprehensively and independently extracted the basic data included in the study, including: first author, data collection time, national and economic level, continent, study design, sample size, age, gender (percentage of males), occupation; numbers of individuals with depression, anxiety, job burnout, post-traumatic stress disorder, and acute stress disorder; and assessment tools used. After the data was extracted, the two reviewers resolved any differences by consensus; if necessary, a third reviewer (J. Huang) was consulted.

### 2.4. Quality assessment

The quality of each study included study in this analysis was evaluated using the American Institute for Healthcare Quality and Research (AHRQ) cross-sectional study evaluation criteria [22] and the Newcastle-Ottawa observational study scale (NOS) [23]. The AHRQ consists of 11 items, each with a score of 0 or 1; with total scores of ≤ 3, 4-7, and ≥ 8 indicating low, medium quality, and high quality, respectively. The NOS consists of eight items that are used to evaluate cohort and case-control studies; scores range from 0-9, with scores <6 regarded as low quality and those ≥6 as high quality. Three reviewers (J. Huang, Z.T. Huang and X.C. Sun) evaluate the quality of each study independently, with any differences resolved by discussions.

### 2.5. Statistical analysis

The comprehensive prevalence rate of mental health problems among health workers during the COVID-19 pandemic was calculated as the main effect, with the metaprop command in STATA used meta-analysis [24]. The random effect model was used to calculate the estimated comprehensive prevalence of mental health disorders to take into account the expected heterogeneity caused by changes in the characteristics of the study. Variability was assessed using the I^2^ index, with indices of 25%, 50% and 75% indicating low, medium and high heterogeneity, respectively, in the random effects model [25]. Subgroup analyses were performed based on six criteria: time of data collection, whether the country was or was not developed, continent, doctors and nurses, doctors/nurses vs. other healthcare workers, and assessment tools. The data collection time was divided into three groups based on the year of data collection, rather than the year of publication: 2020, 2021, and 2022. Countries around the world were divided into developed and developing countries according to the human development index(HDI) proposed by the United Nations Development Programme[26]. The study of other healthcare workers was abandoned when studying doctors and nurses, followed by the scale used by the study. Except for the three subgroups of data collection time, continent and evaluation tool, the other subgroups were defined as binary. The six models were evaluated independently, with each factor examined separately. All statistical analyses were performed using STATA16.0 software, with a two-side p-value <0.05 defined as statistically significant.

## 3. Results

### 3.1. Search results

The literature search identified 7,259 citations, of which 1,595 were deleted due to repetition, and 5,444 were excluded after reading their titles and abstracts. A total of 220 articles were evaluated and screened. Among them, 8 studies were excluded due to topic mismatch, 42 studies were excluded due to inconsistent results, 1 study was excluded as it was not an original research, and 8 studies were excluded as they were letters to journal editors. Detailed reasons for the exclusions can be found in **S2 Table**. Hence, a total of 161 studies were incorporated into this systematic review and meta-analysis, and all of them are appropriately cited in the reference section [27–187]. **Fig 1** shows the flow diagram of the search process and study selection, following the PRISMA protocol.

**Fig 1.**
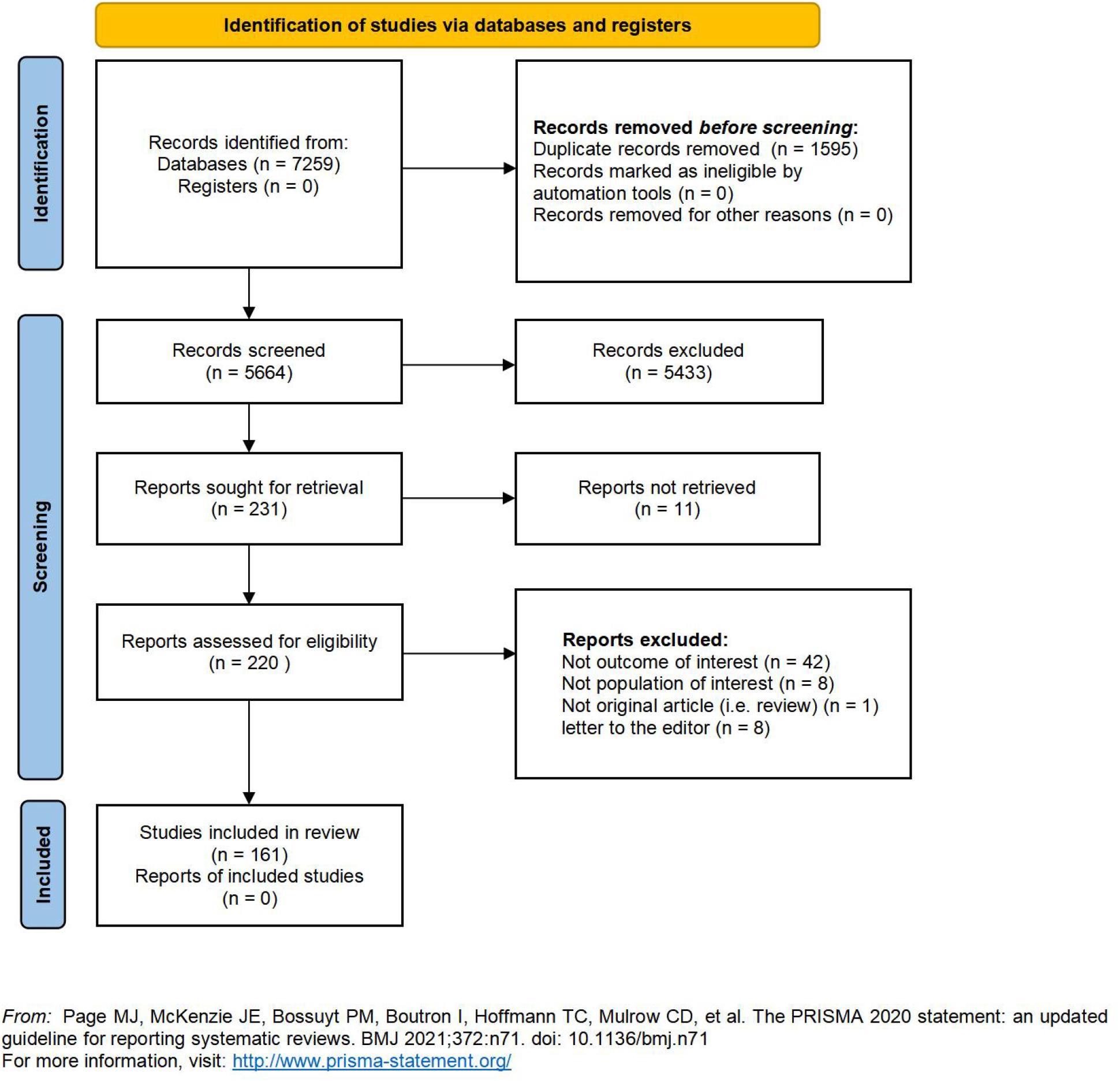
Prisma flow-diagram.

### 3.2. Study characteristics

All 161 included studies focused on the mental health of health workers during the pandemic; of these, 136 studies (84.5%) evaluated anxiety, 134 (83.2%) evaluated depression, 18 (11.2%) evaluated job burnout, 45 (27.9%) evaluated post-traumatic stress disorder, and two (1.2%) evaluated acute stress disorder. Of the 161 included studies, 158 (98.1%) were cross-sectional and three (1.9%) were cohort studies. The studies evaluated healthcare workers from 49 countries on five continents, of which 94 (58.4%), 37 (23.0%), 11 (6.8%), 10 (6.2%), 5 (3.1%) and 4 (2.5%) studies were conducted in Asia, Europe, Africa, North America, South America and Oceania, respectively. The highest number of studies, totaling 34 (21.1%), originated from China, followed by 10 studies (6.2%) from Italy. The United States and Iran tied for third place, each with 8 studies (5.0%). Additionally, three studies (1.9%) were conducted across multiple countries. The 161 studies included 341,014 participants, 82.8% of whom were women. Although not all studies reported age, those that reported age found that participants ranged in age from 18 to ≥60 years. Evaluation of their occupations showed that 16.2% of the participants were doctors, 63.6% were nurses, and 13.3% were other medical personnel. Data for 138 studies (85.7%) were collected in 2020, whereas data for 15 (9.3%) and two (1.2%) studies were collected in 2021 and 2022, respectively, with data collection time not reported for the remaining six (3.7%) studies. Of the 155 studies reporting data collection time, 138 (89%) were collected in 2020, 15 (9.7%) were collected in 2021, and 2 (1.3%) were collected in 2022. **S3, S4 and S5 Tables** present detailed information and quality assessment scores of all included studies.

### 3.3. Meta-analysis of mental health problems among healthcare workers during the COVID-19 pandemic

#### 3.3.1. Burnout

Nineteen studies, including 165,770 participants, reported the prevalence of job burnout, with the combined prevalence rate of job burnout in health workers being 47% (95% CI, 38%-55%, **Fig 2**). Individual study estimates ranged from 12% to 73% and there was evidence of high between-study heterogeneity (I2=99.81%, p<0.001).

**Fig 2.**
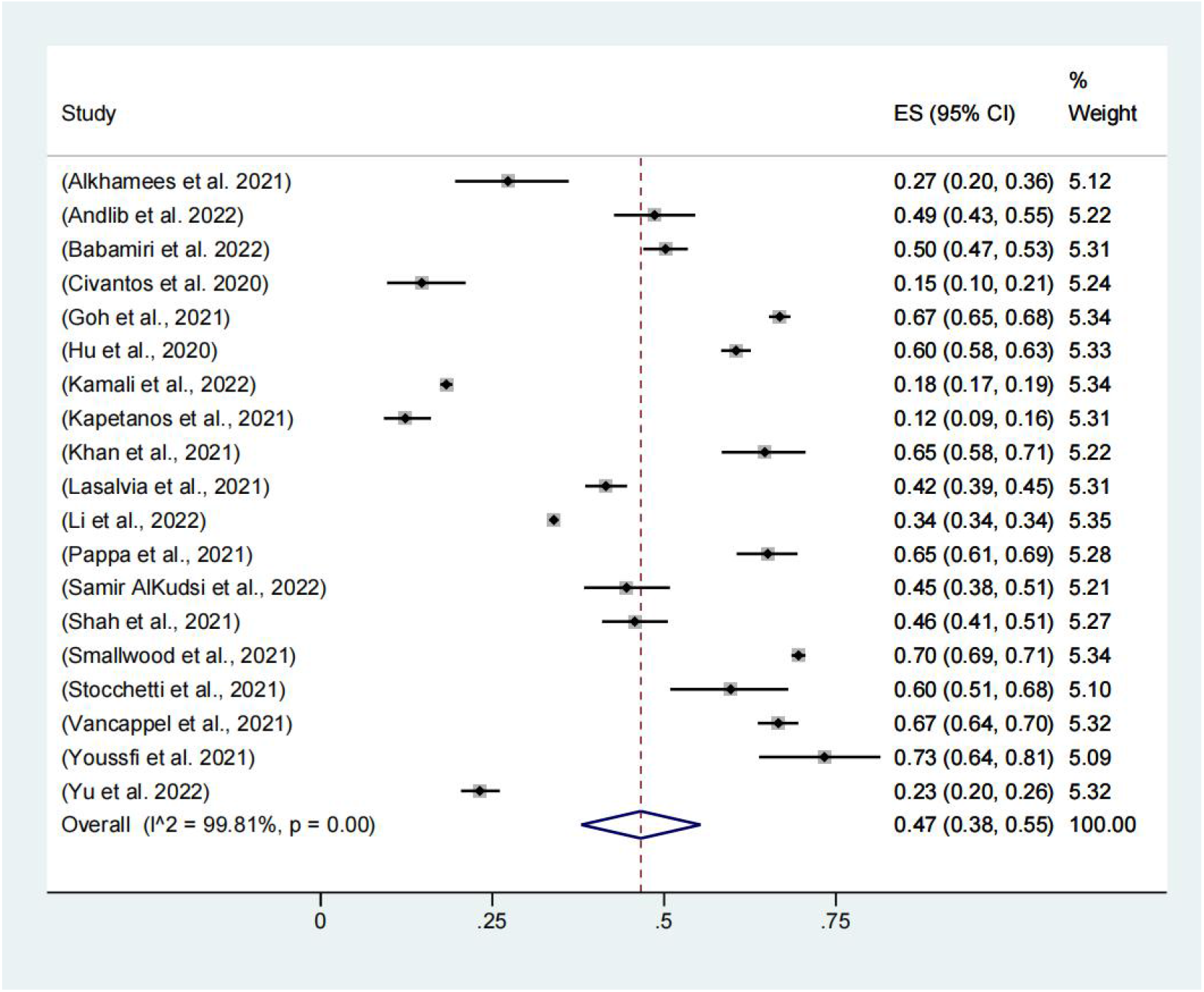
Meta-analysis and pooled estimate of burnout in health care workers during the COVID-19 pandemic.

Subgroup analysis: burnout. As shown in **Table 1**, the combined estimate for 2020 was 47% (95% CI, 33%-61%), and the combined estimate for 2021 was 44% (95% CI, 35%-53%). There was no significant difference in prevalence among different years (p=0.731).

**Table 1.** Subgroup analyses for studies on burnout.

| Subgroup analysis |  | No. of studies | Prevalence %<br>(95% CI) | I<sup>2</sup> | Between-group<br>difference |
| --- | --- | --- | --- | --- | --- |
| Data collection<br>time | 2020 | 15 | 47 (33-61) | 99.82% | p=0.731 |
|  | 2021 | 3 | 44 (35-53) | - |  |
| Continent | Oceania | 1 | 70 (69-71) | - | p<0.001 |
|  | Europe | 5 | 60 (50-70) | - |  |
|  | Africa | 2 | 52 (48-56) | - |  |
|  | Asia | 8 | 37 (28-46) | 99.67% |  |
|  | North America | 2 | 31 (28-33) | - |  |
|  | South America | 1 | 15 (10-21) | - |  |
| Assessment tools | OLBI | 1 | 67 (65-68) | - | p<0.001 |
|  | MBI | 14 | 47 (37-57) | 99.83% |  |
|  | SPFI | 1 | 46 (41-51) | - |  |
|  | ProQoL | 2 | 28 (25-31) | - |  |
|  | Mini-Z | 1 | 15 (10-21) | - |  |

Abbreviation: MBI=Maslach Burnout Inventory; Mini-Z=a non-proprietary, single-item burnout measure; OLBI=Oldenburg Burnout Inventory; ProQoL=Professional Quality of Life scale; SPFI=Stanford Professional Fulfillment Index Questionnaire

It was estimated that the prevalence rate of job burnout was significantly different in different regions (p<0.001). Studies from Oceania had the highest estimated combined prevalence rate (70%; 95% CI, 69%-71%) and the lowest in South America (15%; 95% CI, 10%-21%).

Fourteen studies used the Maslach Job Burnout scale (MBI) [188], and the combined prevalence of all these studies was estimated at 47% (95% CI, 37%-57%). The highest combined prevalence rate was calculated from the study using the Oldenburg Burnout Inventory (OLBI) [189] (67%; 95% CI, 65%-68%), and the study using the single-item Mini-Z burnout assessment [190] obtained the lowest comprehensive estimate (15%; 95% CI, 10%-21%). The combined estimated values of these subgroups were significantly different (p<0.001).

There was no evidence of differential prevalence estimates across other subgroups: the degree of national development(p=0.515); doctors and nurses (p=0.417); and doctors and nurses vs other healthcare workers (p=0.079). **S1-S6 Figs** display the specific results of subgroup analyses.

#### 3.3.2. Anxiety

A total of 136 studies, involving 112,805 participants, reported the prevalence of anxiety disorders, with the combined prevalence rate of anxiety among health workers being 38% (95% CI, 35-41%, **Fig 3**). Individual study estimates ranged from 6% to 90% and there was evidence of high heterogeneity between studies (I2=99.67%, p<0.001).

**Fig 3.**
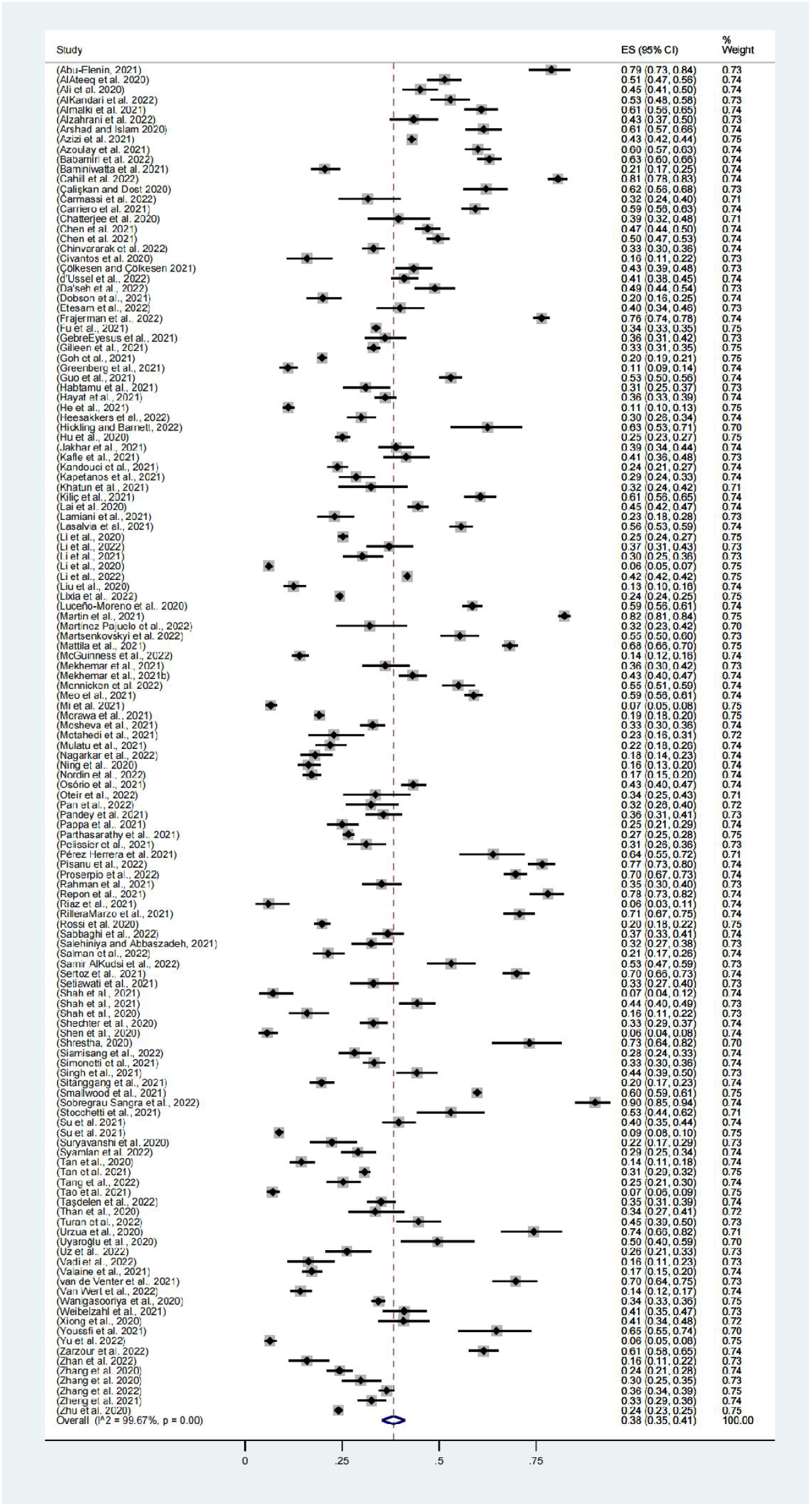
Meta-analysis and pooled estimate of anxiety in health care workers during the COVID-19 pandemic.

Subgroup analysis: anxiety. As shown in **Table 2**, the combined estimate for 2020 was 38% (95% CI, 35%-42%), the combined estimate for 2021 was 41% (95% CI, 34%-47%), and the combined estimate for 2022 was 29% (95% CI, 26%-33%). The prevalence rate of 2022 was lower than that of 2021, and there were significant differences in different years (p<0.001).

**Table 2.** Subgroup analyses for studies on anxiety.

| Subgroup analysis |  | No. of studies | Prevalence %<br>(95% CI) | I<sup>2</sup> | Between-group<br>difference |
| --- | --- | --- | --- | --- | --- |
| Data collection time | 2020 | 117 | 38 (35-42) | 99.60% | p<0.001 |
|  | 2021 | 14 | 41 (34-47) | 98.80% |  |
|  | 2022 | 2 | 29 (26-33) | - |  |
| Assessment tools | STAI | 6 | 56 (39-73) | 99.08% | p<0.001 |
|  | GHQ-28 | 2 | 55 (52-58) | - |  |
|  | PROMIS | 2 | 53 (51-55) | - |  |
|  | GAD-7/BAI | 1 | 50 (40-59) | - |  |
|  | HADS | 19 | 43 (32-55) | 99.67% |  |
|  | ISR | 1 | 41 (35-47) | - |  |
|  | GAD-7 | 54 | 39 (34-45) | 99.73% |  |
|  | DASS-42 | 4 | 38 (28-47) | 92.05% |  |
|  | DASS-21 | 27 | 36 (30-41) | 99.04% |  |
|  | GAD-2 | 4 | 29 (22-36) | 97.45% |  |
|  | SAS | 9 | 25 (14-37) | 99.69% |  |
|  | PHQ-4 | 2 | 18 (17-19) | - |  |
|  | PHQ-2 | 1 | 16 (11-22) | - |  |
|  | CAS | 2 | 11 (10-13) | - |  |
|  | SCL-90 | 2 | 10 (8-12) | - |  |

Abbreviation: BAI=Beck Anxiety Inventory; CAS=Coronavirus Anxiety Scale; DASS-21=The Depression Anxiety Stress Scale-21; DASS-42=The Depression Anxiety Stress Scale-42; GAD-2=The 2-item Generalized Anxiety Disorder Scale; GAD-7=General Anxiety Disorder-7; GHQ-28=General Health Questionnaire; HADS=The Hospital Anxiety and Depression Scale; ISR=The self-report questionnaire ICD-10 Symptom Rating; PROMIS=National Institution of Mental Health (NIMH) Patient-Reported Outcomes Measurement Information System; SAS=Self-Rating Anxiety Scale; SCL-90=Symptom Checklist 90; STAI=State Trait Anxiety Inventory Scale; PHQ-2=The 2-item Generalized Anxiety Disorder Scale; PHQ-4=Patient Health Questionnaire-4

A total of 54 studies used the Generalized Anxiety Disorder 7 (GAD-7) questionnaire [191], and the combined prevalence rate of all these studies was estimated at 39% (95% CI, 34%-45%). The highest combined prevalence rate was calculated from the study using the State-Trait Anxiety Inventory (STAI) [192] (56%; CI, 39%-73%), and the study using Symptom Checklist 90 (SCL-90) [193] produced the lowest comprehensive estimate (10%; 95% CI, 8%-12%). The combined estimated values of these subgroups were significantly different (p<0.001).

There was no evidence of differential prevalence estimates across other subgroups: the degree of national development(p=0.435); continents (p=0.298); doctor and nurse (p=0.796); and doctors and nurses vs other healthcare workers (p=0.378). **S7-S12 Figs** display the specific results of subgroup analyses.

#### 3.3.3. Depression

A total of 134 studies, involving 316,891 participants, reported the prevalence of depression, with the combined prevalence rate of depression among healthcare workers being 34% (95% CI, 30-38%, **Fig 4**). Estimates for a single study ranged from 4% to 91%, and there was evidence of high heterogeneity between studies (I2=99.81%, p<0.001).

**Fig 4.**
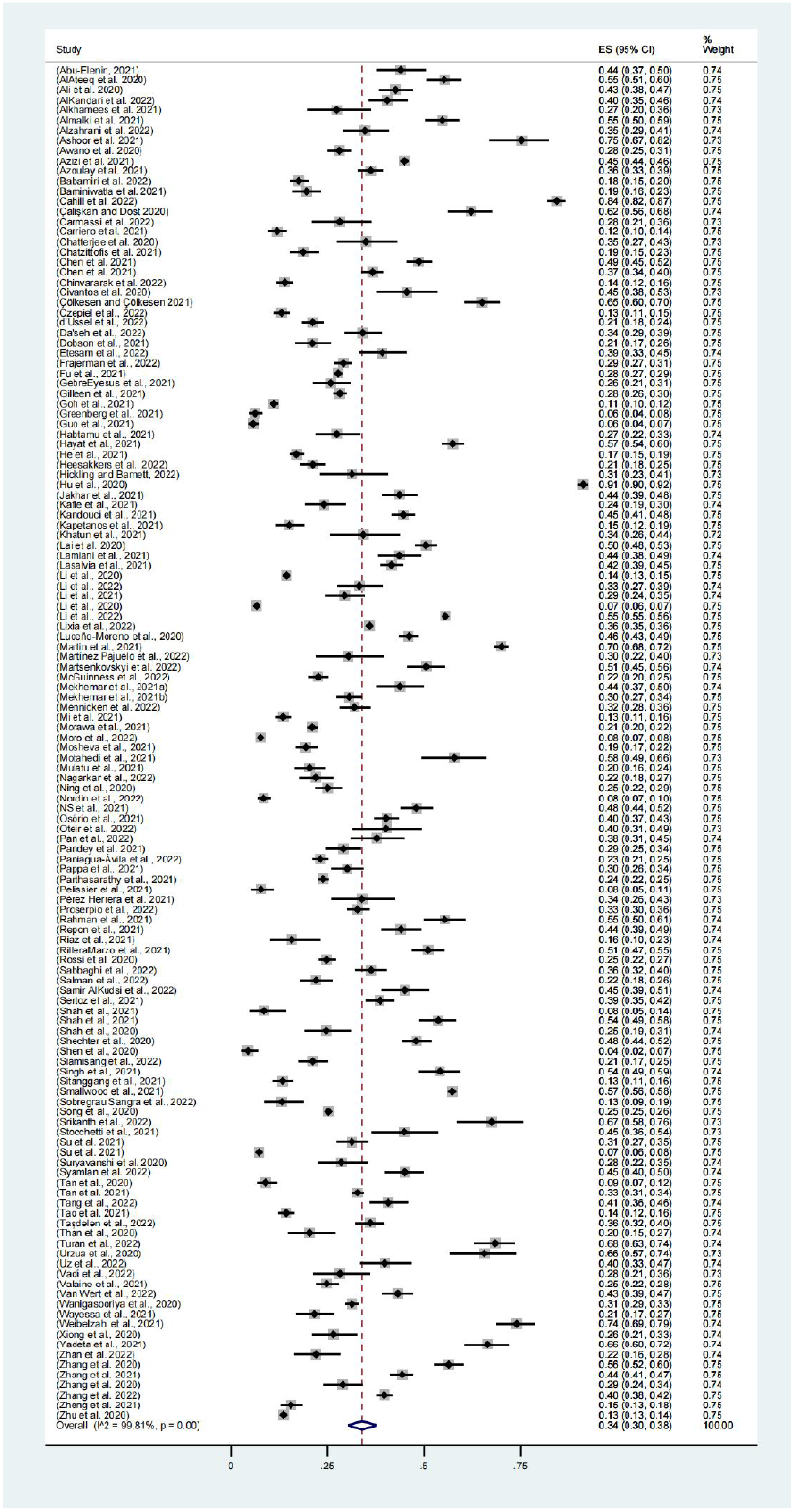
Meta-analysis and pooled estimate of depression in health care workers during the COVID-19 pandemic.

Subgroup analysis: depression. As shown in **Table 3**, the combined estimate for 2020 was 33% (95% CI, 30%-37%), the combined estimate for 2021 was 38% (95% CI, 27%-48%), and the combined estimate for 2022 was 38% (95% CI, 34%-41%). The prevalence rate in 2021 and 2022 was higher than that in 2020, although statistical analysis showed that there was no significant difference in prevalence among different years (p=0.255).

**Table 3.**
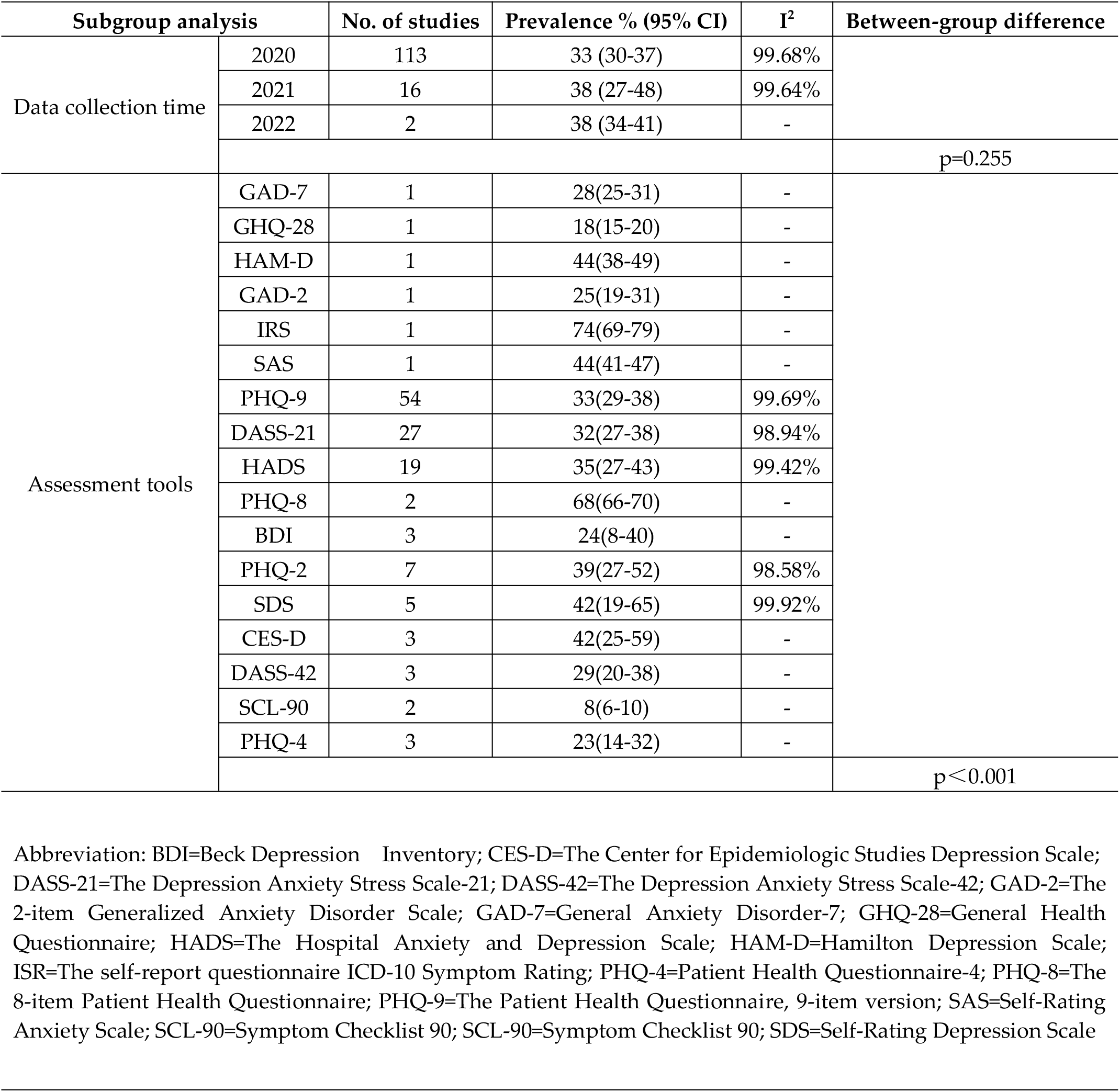
Subgroup analyses for studies on depression.

Fifty-four studies used the patient Health questionnaire (PHQ-9) [194], and the combined prevalence rate of all these studies was estimated at 33% (95% CI, 29%-38%). The highest combined prevalence rate was calculated from the study using the ICD-10 symptom Checklist 90 (ISR) [195,196] (74%; 95% CI, 69%-79%), while the study using the symptom list 90 (SCL-90) [193] yielded the lowest comprehensive estimate (8%; 95% CI, 6%-10%). The combined estimated values of these subgroups were significantly different (p<0.001).

There was no evidence of differential prevalence estimates across other subgroups: the degree of national development(p=0.321); continents (p=0.352); doctor and nurse (p=0.746); and doctors and nurses vs other healthcare workers (p=0.196). **S13-S18 Figs** display the specific results of subgroup analyses.

#### 3.3.4. Acute stress disorder

Two studies, involving 5,776 participants, reported the prevalence of acute stress disorder, with the combined prevalence rate of acute stress disorder among healthcare workers being 30% (95% CI, 29-31%, **Fig 5**). The data were collected in 2020. Two evaluation scales were used: Standford acute Stress Reaction Questionnaire (SARS-Q) (197) and Impact of Event Scale-Revised Questionnaires (IES-R) (198).

**Fig 5.**
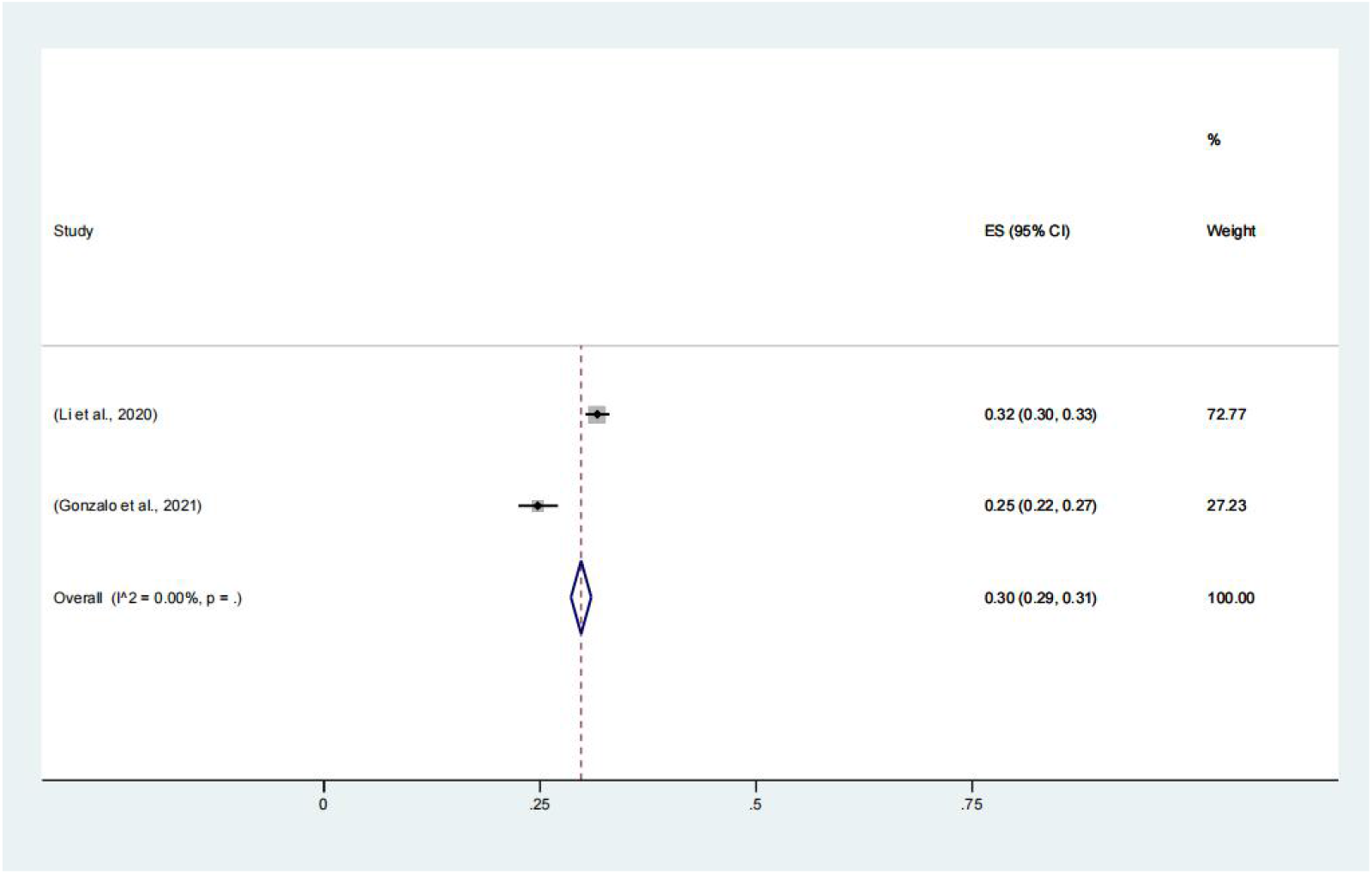
Meta-analysis and pooled estimate of acute stress disorder in health care workers during the COVID-19 pandemic.

No subgroup analysis was performed because few studies reported the prevalence of acute stress disorder (<3).

#### 3.3.5. Post-traumatic stress disorder

Forty-four studies, including 89,977 participants, reported the prevalence of post-traumatic stress disorder (PTSD), with the combined prevalence rate of PTSD among healthcare workers being 26% (95% CI, 22-29%, **Fig 6**).Individual study estimates ranged from 2% to 67% and there was evidence of high between-study heterogeneity (I2=99.64%, p<0.001).

**Fig 6.**
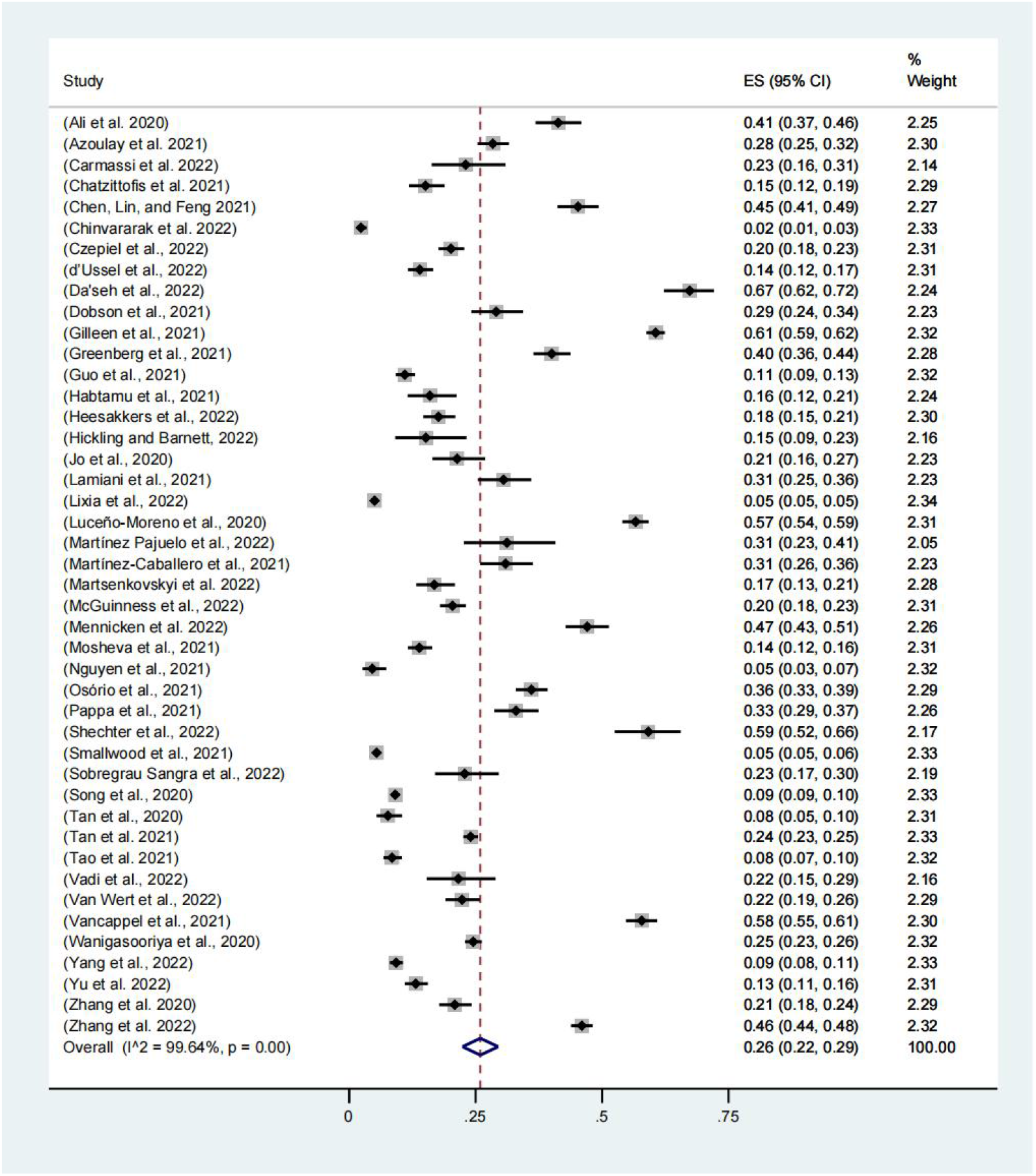
Meta-analysis and pooled estimate of post-traumatic stress disorder in health care workers during the COVID-19 pandemic.

Subgroup analysis: post-traumatic stress disorder. As shown in **Table 4**, the combined estimate for 2020 was 25% (95% CI, 21%-29%) and for 2021 was 28% (95% CI, 13%-42%). The prevalence rate was similar in different years and there was no significant difference (p=0.733).

**Table 4.**
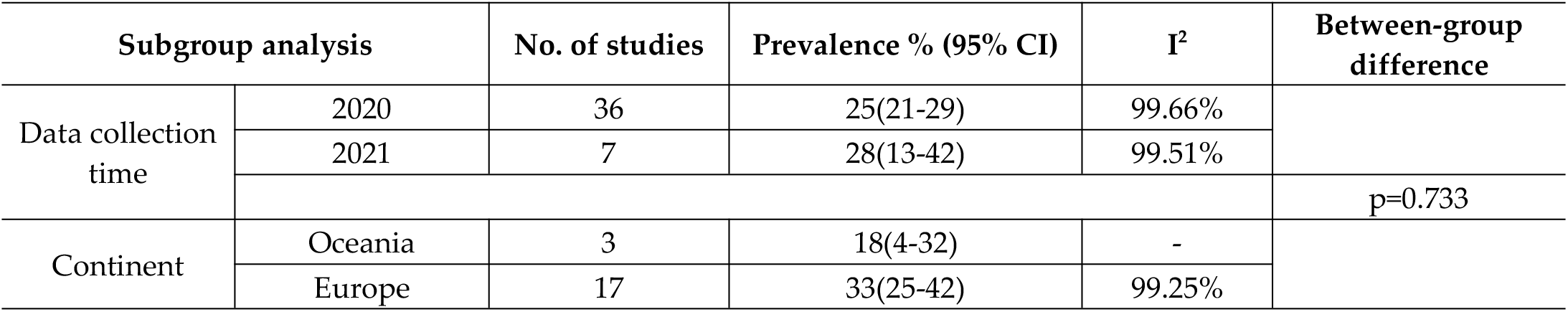
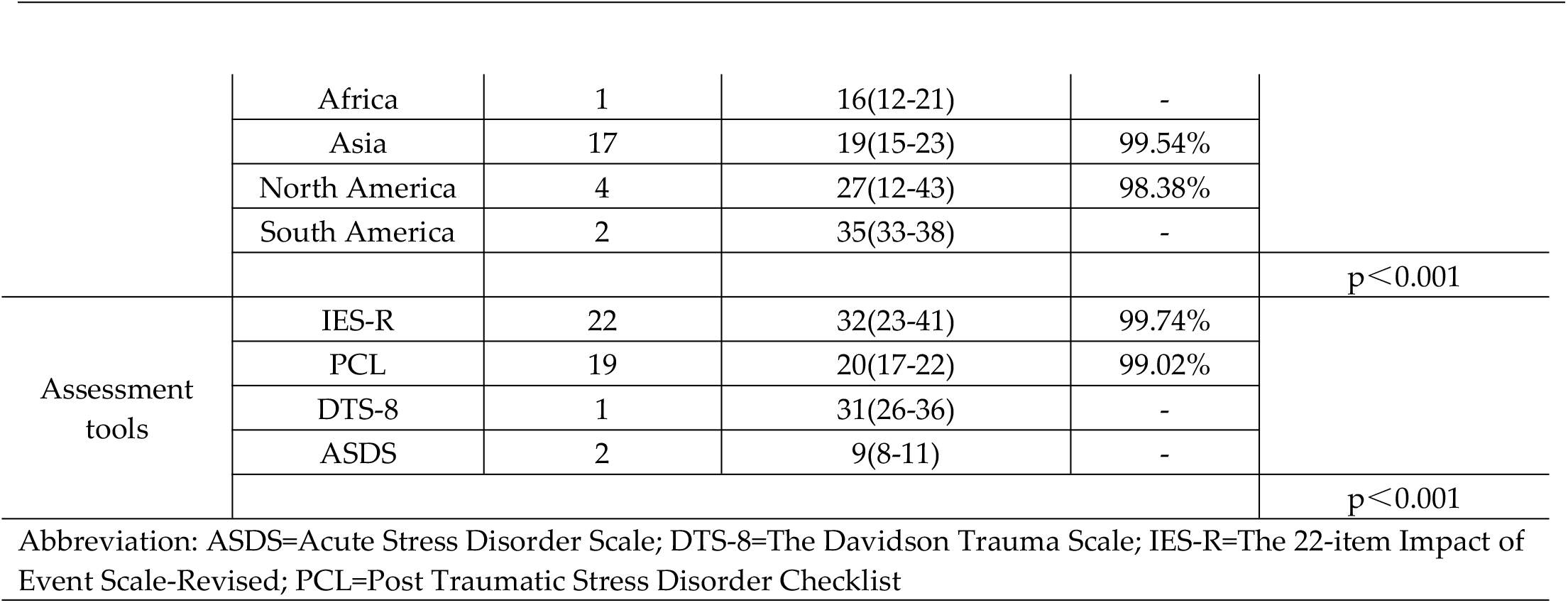
Subgroup analyses for studies on post-traumatic stress disorder.

It was estimated that the prevalence rate of post-traumatic stress disorder was significantly different in different regions (p< 0.001). The study from South America produced the highest estimated combined prevalence rate (35%; 95% CI, 33%-38%) and the lowest in Africa (16%; 95% CI, 12%-21%).

Twenty-two studies used the Impact of Event Scale-Revised (IES-R) [198], and the combined prevalence rate of all these studies was estimated at 32% (95% CI, 23%-41%), which was the highest. The study using the Acute Stress Disorder Scale (ASDS) [199] yielded the lowest comprehensive estimate (9%; 95% CI, 8%-11%). The combined estimated values of these subgroups were significantly different (p<0.001).

There was no evidence of differential prevalence estimates across other subgroups: the degree of national development(p=0.107); doctors and nurses (p=0.336); and doctors and nurses vs other healthcare workers (p=0.473). **S19-S24 Figs** display the specific results of subgroup analyses.

### 3.4. Quality assessment

The quality of the 158 cross-sectional studies was evaluated using the AHRQ [22], which found that the average score was 6.03 ±1.12. Of these 158 studies, 17 (10.8%) had scores ≥ 8, indicating high-quality, and the remaining 141 (89.2%) had scores of 4 to 7, indicating medium-quality; none had scores <4, indicating low quality. Evaluation of the three cohort studies with the NOS scale[23] showed that their average score was 6.67 ±0.47, with all three studies having scores ≥ 6, indicating high quality. **S4 and S5 tables** provides details of the study quality scores.

## 4. Discussion

This meta-analysis of 161 studies involving 341,014 healthcare workers was performed to provide an update on the prevalence of multiple mental health symptoms among healthcare workers worldwide. During the COVID-19 pandemic, the pooled prevalence rates of burnout, anxiety, depression, acute stress disorder and post-traumatic stress disorder among healthcare workers were 47%, 38%, 34%, 30% and 26%, respectively. These rates were higher than in the general population [200–202], suggesting that mental health risks were greater for healthcare workers during the pandemic.

Evaluation of 136 studies showed that the prevalence rate of anxiety among medical workers was 38%, higher than the 26.9% [200] and 20% [201] rates reported in the general population. The 134 studies that assessed depression reported a prevalence among healthcare workers of 34%, higher than that of 28% [200] and 30% [201] rates in the general population. Anxiety and depression among medical staff during COVID-19 were related to many factors, such as the shortage of personal protective equipment, the lack of other medical resources and the sacrifice of colleagues [5]. Most healthcare workers believed that, despite having adequate personal protective equipment, they felt unsafe when contacting and treating patients. Moreover, they were also worried about their families becoming infected, especially if they take care of these infected patients, with the possibility that infection may spread to their families [4, 203]. This pandemic resulted in an unprecedented restructuring of the healthcare system, with a large number of medical staff, especially nurses, being transferred to intensive care units [204]. These workers had to absorb a lot of information and master new technical skills associated with unfamiliar areas of expertise in a short period of time, which could also lead to mental health disorders [205].

Few studies in a previous meta-analysis focused on job burnout of healthcare workers,our analysis showed that the combined prevalence rate of burnout among medical workers in 18 studies was 47%, higher than that of 37% in previous studies [18]. Job burnout among healthcare workers was high even before the pandemic [206]. During the pandemic, burnout was further exacerbated by long-term intense and stressful work, emotional exhaustion, deteriorating mental health, fear of infection and lack of support [207].The increasing death toll of critically ill patients not only led to depression and pain among healthcare workers, but reduced their sense of professional achievement [208]. This not only had a negative impact on the physical and mental health and interpersonal relationships of medical personnel, but also affected their attitudes toward work and their efficiency.In addition, the present meta-analysis showed that the prevalence of acute stress disorder among health care workers was 30%, and the prevalence of PTSD was 26%, rates significantly higher than earlier in the pandemic [209].

When interpreting the pooled prevalence estimates calculated in this review and meta-analysis, it is need to note that the percentage of variability (I^2^) in the prevalence estimates due to heterogeneity was very high.When a large number of studies are included in meta-analyses, the I^2^ is extremely sensitive, while a high I^2^ is often inevitable [210]. The I^2^ may therefore detect only a small amount of heterogeneity, which is not clinically important. Despite this, we discussed the heterogeneity through subgroup analysis.Although the subgroup analyses suggested evidence of between-group differences in prevalence estimates across a number of variables (e.g. data collection time, continent, evaluation tool), intra-group heterogeneity is still high and does not fully explain the existence of heterogeneity.

Our subgroup analysis found that the prevalence of anxiety remained at a high level in 2020 and 2021, and showed a significant downward trend in 2022. The prevalence rates of job burnout, depression, and post-traumatic stress disorder (PTSD) had consistently remained high, and there was an increasing trend in depression and PTSD, which was consistent with previous research conducted on SARS [211]. PTSD has been evaluated infrequently in previous studies, as it usually occurs weeks or months after the acute stress disorder; that is, it appears later in the “repair phase” after the virus has been roughly controlled. In addition to having adverse psychological consequences, PTSD can lead to changes in a variety of physiological functions and have deleterious effects on multiple organ systems, especially increasing the risk of cardiovascular disease. Moreover, the presence of post-traumatic stress disorder (PTSD) among healthcare workers could potentially have detrimental effects on the healthcare system. Therefore, we suggest implementing screening measures and interventions for healthcare personnel [212,213]. The decline in the prevalence of anxiety may be due to the rapid and positive response of all countries and regions around the world after the pandemic, including expanding the production of medical supplies and calling on citizens to wear masks [214].The major breakthrough in novel coronavirus’s research has led to the successful research and development of vaccines [215] and the advent of drugs aimed at novel coronavirus [216], as well as the accumulated experience of medical staff and broad social support, which quickly stabilized the tense situation and reduced the negative impact of the pandemic.

However, it should be noted that the pandemic is not really over [217], and the continuous emergence of new virulent strains such as AY.4.2 [218] and XBB.1.5 [219], as well as changes in governments’ pandemic prevention and control policies, may cause a new round of outbreaks at any time. Repeated psychological damage to health care workers will not only increase the risk of illness and lead to the deterioration of psychological disorders, but also lead to an increase in the prevalence and death of serious diseases [213]. Mental health disorders of medical staff are not as easy to detect as physical injuries, but they can also cause great pain to medical staff and damage the health care system and public safety[211]. Therefore, the prevention and control of the pandemic should not be taken lightly, and global leaders and all stakeholders should devote resources to the mental health of health care workers to screen for mental health problems and take effective interventions to help them solve their mental health disorders.

In addition, this study also has some limitations. Although seven major databases were searched, some of the other unsearched databases were still omitted and the search was mainly focused on English literature, and there were no grey literature. Secondly, during the pandemic, in order to clarify the biological characteristics of COVID-19 virus and develop better diagnosis and treatment strategies, a large number of studies related to novel coronavirus were carried out and published [220], but the quality of some of them was limited [221]. And most of the studies on the prevalence of mental disorders are cross-sectional studies, and the lower level of evidence in evidence-based medicine affects the credibility of the study [222].Because different pandemic stages of each study lead to higher heterogeneity in the estimated prevalence rate, and different pandemic stages in different countries, there is no very ideal indicator to define different periods of the global epidemic. These also affect the accuracy of the survey results.

## 5. Conclusions

Following the outbreak of the COVID-19 pandemic, the prevalence rates of mental health disorder among healthcare workers worldwide increased significantly, resulting in serious damage to their physical and mental health. Although the pandemic has been brought under control, it has not really disappeared. Thus, future outbreaks may further aggravate mental health disorders in healthcare workers. In addition, later-occurring mental health disorders such as PTSD can have serious adverse consequences. Governments and relevant organizations should pay close attention to the mental health status of healthcare workers and implement intervention measures. Moreover, additional research is needed to better prevent and reduce mental health disorders among healthcare workers.

## Supporting information

**S1 File Retrieval strategy.**

**S2 File PRISMA statement and checklist (S1 Table).**

**S3 File Reasons for exclusion during full-text screening (S2 Table).**

**S4 File Main characteristics of the included studies (S3 Table).**

**S5 File Quality assessment (S4 and S5 Table).**

**S6 File Other subgroup analysis results (S1-S24 Figs).**

## Author Contributions

**Conceptualization:** J. Huang.

**Methodology:** J. Huang.

**Software:** J. Huang.

**Validation:** J. Huang., X.C. Sun. and Z.T. Huang.

**Formal analysis:** J. Huang.

**Investigation:** J. Huang., X.C. Sun. and Z.T. Huang.

**Resources:** J. Huang. and T.T.Chen.

**Data curation:** J. Huang. and Z.T. Huang.

**Writing—original draft preparation:** J. Huang. **Writing—review and editing:** J. Huang. and X.T Wu.

**Visualization**: J. Huang. and Z.T. Huang.

**Supervision:** J. Huang.

**Project administration:** J. Huang.

**Funding acquisition:** J. Huang.

**All authors have read and agreed to the published version of the manuscript.**

## Data Availability Statement

The data used in this meta-analysis are derived from previously published studies and are publicly available. The datasets analyzed during the current study are available in the references cited.

## Data citation

Please refer to the respective publications for the datasets used in this meta-analysis.

## Acknowledgments

The authors thank all the individuals who participated in this study.

## References

1. 1. World Health Organization. (2020, January 9). WHO Statement Regarding Cluster of Pneumonia Cases in Wuhan, China. Retrieved January 11, 2020, from https://www.who.int/china/news/detail/09-01-2020-who-statement-regarding-cluster-of-pneumonia-cases-in-wuhan-china

2. World Health Organization. (2023). WHO Coronavirus COVID-19 Dashboard | WHO Coronavirus COVID-19 Dashboard With Vaccination Data. Retrieved from https://covid19.who.int/

3. Vignaud P, Prieto N. [Psychological impact of the COVID-19 pandemic on health care professionals]. Actual Pharm. 2020;59(599):51–3. Epub 2020/08/31. doi: 10.1016/j.actpha.2020.08.013. PubMed PMID: 32863554; PubMed Central PMCID: PMCPMC7441863.

4. Goulia P, Mantas C, Dimitroula D, Mantis D, Hyphantis T. General hospital staff worries, perceived sufficiency of information and associated psychological distress during the A/H1N1 influenza pandemic. BMC infectious diseases. 2010;10:322. Epub 2010/11/11. doi: 10.1186/1471-2334-10-322. PubMed PMID: 21062471; PubMed Central PMCID: PMCPMC2990753.

5. Hall H. The effect of the COVID-19 pandemic on healthcare workers’ mental health. JAAPA : official journal of the American Academy of Physician Assistants. 2020;33(7):45–8. Epub 2020/06/27. doi: 10.1097/01.JAA.0000669772.78848.8c. PubMed PMID: 32590533.

6. Santolalla-Arnedo I, Pozo-Herce PD, De Viñaspre-Hernandez RR, Gea-Caballero V, Juarez-Vela R, Gil-Fernandez G, et al. Psychological impact on care professionals due to the SARS-Cov-2 virus in Spain. International nursing review. 2022;69(4):520–8. Epub 2022/02/03. doi: 10.1111/inr.12748. PubMed PMID: 35107171; PubMed Central PMCID: PMCPMC9790592.

7. Del Pozo-Herce P, Garrido-García R, Santolalla-Arnedo I, Gea-Caballero V, García-Molina P, Ruiz de Viñaspre-Hernández R, et al. Psychological Impact on the Nursing Professionals of the Rioja Health Service (Spain) Due to the SARS-CoV-2 Virus. International journal of environmental research and public health. 2021;18(2). Epub 2021/01/16. doi: 10.3390/ijerph18020580. PubMed PMID: 33445563; PubMed Central PMCID: PMCPMC7827934.

8. Chong MY, Wang WC, Hsieh WC, Lee CY, Chiu NM, Yeh WC, et al. Psychological impact of severe acute respiratory syndrome on health workers in a tertiary hospital. The British journal of psychiatry : the journal of mental science. 2004;185:127–33. Epub 2004/08/03. doi: 10.1192/bjp.185.2.127. PubMed PMID: 15286063.

9. Lee AM, Wong JG, McAlonan GM, Cheung V, Cheung C, Sham PC, et al. Stress and psychological distress among SARS survivors 1 year after the outbreak. Canadian journal of psychiatry Revue canadienne de psychiatrie. 2007;52(4):233–40. Epub 2007/05/16. doi: 10.1177/070674370705200405. PubMed PMID: 17500304.

10. Anjum S, Ullah R, Rana MS, Khan HA, Memon FS, Ahmed Y, et al. COVID-19 Pandemic: A Serious Threat for Public Mental Health Globally. Psychiatria Danubina. 2020;32(2):245–50. Epub 2020/08/17. doi: 10.24869/psyd.2020.245. PubMed PMID: 32796793.

11. Xiong J, Lipsitz O, Nasri F, Lui LMW, Gill H, Phan L, et al. Impact of COVID-19 pandemic on mental health in the general population: A systematic review. Journal of affective disorders. 2020;277:55–64. Epub 2020/08/18. doi: 10.1016/j.jad.2020.08.001. PubMed PMID: 32799105; PubMed Central PMCID: PMCPMC7413844.

12. Bohórquez-Blanco S, Allande-Cussó R, Martín-López C, Gómez-Salgado J, García-Iglesias JJ, Fagundo-Rivera J, et al. Effects of the COVID-19 pandemic on the mental health of rehabilitation area professionals: A systematic review. Frontiers in public health. 2022;10:1085820. Epub 2022/12/27. doi: 10.3389/fpubh.2022.1085820. PubMed PMID: 36568762; PubMed Central PMCID: PMCPMC9779931.

13. Gourret Baumgart J, Kane H, El-Hage W, Deloyer J, Maes C, Lebas MC, et al. The Early Impacts of the COVID-19 Pandemic on Mental Health Facilities and Psychiatric Professionals. International journal of environmental research and public health. 2021;18(15). Epub 2021/08/08. doi: 10.3390/ijerph18158034. PubMed PMID: 34360334; PubMed Central PMCID: PMCPMC8345711.

14. Paiano M, Jaques AE, Nacamura PAB, Salci MA, Radovanovic CAT, Carreira L. Mental health of healthcare professionals in China during the new coronavirus pandemic: an integrative review. Revista brasileira de enfermagem. 2020;73(suppl 2):e20200338. Epub 2020/09/24. doi: 10.1590/0034-7167-2020-0338. PubMed PMID: 32965402.

15. Deng J, Zhou F, Hou W, Silver Z, Wong CY, Chang O, et al. The prevalence of depression, anxiety, and sleep disturbances in COVID-19 patients: a meta-analysis. Annals of the New York Academy of Sciences. 2021;1486(1):90–111. Epub 2020/10/04. doi: 10.1111/nyas.14506. PubMed PMID: 33009668; PubMed Central PMCID: PMCPMC7675607.

16. Hill JE, Harris C, Danielle LC, Boland P, Doherty AJ, Benedetto V, et al. The prevalence of mental health conditions in healthcare workers during and after a pandemic: Systematic review and meta-analysis. Journal of advanced nursing. 2022;78(6):1551–73. Epub 2022/02/13. doi: 10.1111/jan.15175. PubMed PMID: 35150151; PubMed Central PMCID: PMCPMC9111784.

17. Saragih ID, Tonapa SI, Saragih IS, Advani S, Batubara SO, Suarilah I, et al. Global prevalence of mental health problems among healthcare workers during the Covid-19 pandemic: A systematic review and meta-analysis. International journal of nursing studies. 2021;121:104002. Epub 2021/07/17. doi: 10.1016/j.ijnurstu.2021.104002. PubMed PMID: 34271460; PubMed Central PMCID: PMCPMC9701545.

18. Aymerich C, Pedruzo B, Pérez JL, Laborda M, Herrero J, Blanco J, et al. COVID-19 pandemic effects on health worker’s mental health: Systematic review and meta-analysis. European psychiatry : the journal of the Association of European Psychiatrists. 2022;65(1):e10. Epub 2022/01/22. doi: 10.1192/j.eurpsy.2022.1. PubMed PMID: 35060458; PubMed Central PMCID: PMCPMC8828390.

19. Li Y, Scherer N, Felix L, Kuper H. Prevalence of depression, anxiety and post-traumatic stress disorder in health care workers during the COVID-19 pandemic: A systematic review and meta-analysis. PloS one. 2021;16(3):e0246454. Epub 2021/03/11. doi: 10.1371/journal.pone.0246454. PubMed PMID: 33690641; PubMed Central PMCID: PMCPMC7946321.

20. Lee BEC, Ling M, Boyd L, Olsson C, Sheen J. The prevalence of probable mental health disorders among hospital healthcare workers during COVID-19: A systematic review and meta-analysis. Journal of affective disorders. 2023;330:329–45. Epub 2023/03/18. doi: 10.1016/j.jad.2023.03.012. PubMed PMID: 36931567; PubMed Central PMCID: PMCPMC10017178.

21. Moher, D., Liberati, A., Tetzlaff, J., Altman, D. G., & PRISMA Group (2009). Preferred reporting items for systematic reviews and meta-analyses: the PRISMA statement. PLoS medicine, 6(7), e1000097. https://doi.org/10.1371/journal.pmed.1000097

22. Viswanathan M, Ansari MT, Berkman ND, Chang S, Hartling L, McPheeters M, et al. AHRQ Methods for Effective Health Care

23. Assessing the Risk of Bias of Individual Studies in Systematic Reviews of Health Care Interventions. Methods Guide for Effectiveness and Comparative Effectiveness Reviews. Rockville (MD): Agency for Healthcare Research and Quality (US); 2008.

24. Stang A. Critical evaluation of the Newcastle-Ottawa scale for the assessment of the quality of nonrandomized studies in meta-analyses. Eur J Epidemiol. 2010;25(9):603–5. Epub 2010/07/24. doi: 10.1007/s10654-010-9491-z. PubMed PMID: 20652370.

25. Nyaga VN, Arbyn M, Aerts M. Metaprop: a Stata command to perform meta-analysis of binomial data. Archives of public health = Archives belges de sante publique. 2014;72(1):39. Epub 2014/01/01. doi: 10.1186/2049-3258-72-39. PubMed PMID: 25810908; PubMed Central PMCID: PMCPMC4373114.

26. Roser M. Human Development Index (HDI). Our World in Data. 2014. Available from: https://ourworldindata.org/human-development-index

27. Huedo-Medina TB, Sánchez-Meca J, Marín-Martínez F, Botella J. Assessing heterogeneity in meta-analysis: Q statistic or I2 index? Psychological methods. 2006;11(2):193–206. Epub 2006/06/21. doi: 10.1037/1082-989x.11.2.193. PubMed PMID: 16784338.

28. Dolan ED, Mohr D, Lempa M, Joos S, Fihn SD, Nelson KM, et al. Using a single item to measure burnout in primary care staff: a psychometric evaluation. Journal of general internal medicine. 2015;30(5):582–7. Epub 2014/12/03. doi: 10.1007/s11606-014-3112-6. PubMed PMID: 25451989; PubMed Central PMCID: PMCPMC4395610.

29. Spitzer RL, Kroenke K, Williams JB, Löwe B. A brief measure for assessing generalized anxiety disorder: the GAD-7. Archives of internal medicine. 2006;166(10):1092–7. Epub 2006/05/24. doi: 10.1001/archinte.166.10.1092. PubMed PMID: 16717171.

30. Guillén-Riquelme A, Buela-Casal G. [Meta-analysis of group comparison and meta-analysis of reliability generalization of the State-Trait Anxiety Inventory Questionnaire (STAI)]. Revista espanola de salud publica. 2014;88(1):101–12. Epub 2014/04/15. doi: 10.4321/s1135-57272014000100007. PubMed PMID: 24728394.

31. Sereda Y, Dembitskyi S. Validity assessment of the symptom checklist SCL-90-R and shortened versions for the general population in Ukraine. BMC psychiatry. 2016;16(1):300. Epub 2016/08/27. doi: 10.1186/s12888-016-1014-3. PubMed PMID: 27561313; PubMed Central PMCID: PMCPMC5000449.

32. Kroenke K, Spitzer RL, Williams JB. The PHQ-9: validity of a brief depression severity measure. Journal of general internal medicine. 2001;16(9):606–13. Epub 2001/09/15. doi: 10.1046/j.1525-1497.2001.016009606.x. PubMed PMID: 11556941; PubMed Central PMCID: PMCPMC1495268.

33. Tritt K, von Heymann F, Zaudig M, Zacharias I, Söllner W, Loew T. [Development of the “ICD-10-Symptom-Rating”(ISR) questionnaire]. Zeitschrift fur Psychosomatische Medizin und Psychotherapie. 2008;54(4):409–18. Epub 2008/12/04. doi: 10.13109/zptm.2008.54.4.409. PubMed PMID: 19049690.

34. Fischer HF, Tritt K, Klapp BF, Fliege H. [Factor structure and psychometric properties of the ICD-10-Symptom-Rating (ISR) in samples of psychosomatic patients]. Psychotherapie, Psychosomatik, medizinische Psychologie. 2010;60(8):307–15. Epub 2009/04/29. doi: 10.1055/s-0029-1214419. PubMed PMID: 19399700.

35. Cardeña E, Koopman C, Classen C, Waelde LC, Spiegel D. Psychometric properties of the Stanford Acute Stress Reaction Questionnaire (SASRQ): a valid and reliable measure of acute stress. J Trauma Stress. 2000;13(4):719–34. Epub 2000/01/11. doi: 10.1023/a:1007822603186. PubMed PMID: 11109242.

36. Creamer M, Bell R, Failla S. Psychometric properties of the Impact of Event Scale - Revised. Behav Res Ther. 2003;41(12):1489–96. Epub 2004/01/07. doi: 10.1016/j.brat.2003.07.010. PubMed PMID: 14705607.

37. Wang R, Wang L, Zhang J, Liu Z, Wu K. The structure of acute stress disorder among Chinese adults exposed to an earthquake: is dysphoric arousal a unique construct of acute posttraumatic responses? Scandinavian journal of psychology. 2012;53(5):430–6. Epub 2012/08/14. doi: 10.1111/j.1467-9450.2012.00965.x. PubMed PMID: 22882702.

38. Nochaiwong S, Ruengorn C, Thavorn K, Hutton B, Awiphan R, Phosuya C, et al. Global prevalence of mental health issues among the general population during the coronavirus disease-2019 pandemic: a systematic review and meta-analysis. Scientific reports. 2021;11(1):10173. Epub 2021/05/15. doi: 10.1038/s41598-021-89700-8. PubMed PMID: 33986414; PubMed Central PMCID: PMCPMC8119461.

39. Alzahrani F, Alshahrani NZ, Abu Sabah A, Zarbah A, Abu Sabah S, Mamun MA. Prevalence and factors associated with mental health problems in Saudi general population during the coronavirus disease 2019 pandemic: A systematic review and meta-analysis. PsyCh journal. 2022;11(1):18–29. Epub 2022/01/06. doi: 10.1002/pchj.516. PubMed PMID: 34986503.

40. Shi L, Lu ZA, Que JY, Huang XL, Liu L, Ran MS, et al. Prevalence of and Risk Factors Associated With Mental Health Symptoms Among the General Population in China During the Coronavirus Disease 2019 Pandemic. JAMA network open. 2020;3(7):e2014053. Epub 2020/07/02. doi: 10.1001/jamanetworkopen.2020.14053. PubMed PMID: 32609353; PubMed Central PMCID: PMCPMC7330717.

41. Usul E, Şan I, Bekgöz B. The Effect of the COVID-19 Pandemic on the Anxiety Level of Emergency Medical Services Professionals. Psychiatria Danubina. 2020;32(3-4):563–9. Epub 2020/12/29. doi: 10.24869/psyd.2020.563. PubMed PMID: 33370767.

42. El-Hage W, Hingray C, Lemogne C, Yrondi A, Brunault P, Bienvenu T, et al. [Health professionals facing the coronavirus disease 2019 (COVID-19) pandemic: What are the mental health risks?]. L’Encephale. 2020;46(3s):S73–s80. Epub 2020/05/07. doi: 10.1016/j.encep.2020.04.008. PubMed PMID: 32370984; PubMed Central PMCID: PMCPMC7174182.

43. Zhu N, Zhang D, Wang W, Li X, Yang B, Song J, et al. A Novel Coronavirus from Patients with Pneumonia in China, 2019. The New England journal of medicine. 2020;382(8):727–33. Epub 2020/01/25. doi: 10.1056/NEJMoa2001017. PubMed PMID: 31978945; PubMed Central PMCID: PMCPMC7092803.

44. Rodrigues H, Cobucci R, Oliveira A, Cabral JV, Medeiros L, Gurgel K, et al. Burnout syndrome among medical residents: A systematic review and meta-analysis. PloS one. 2018;13(11):e0206840. Epub 2018/11/13. doi: 10.1371/journal.pone.0206840. PubMed PMID: 30418984; PubMed Central PMCID: PMCPMC6231624.

45. Giusti EM, Pedroli E, D’Aniello GE, Stramba Badiale C, Pietrabissa G, Manna C, et al. The Psychological Impact of the COVID-19 Outbreak on Health Professionals: A Cross-Sectional Study. Front Psychol. 2020;11:1684. Epub 2020/08/06. doi: 10.3389/fpsyg.2020.01684. PubMed PMID: 32754102; PubMed Central PMCID: PMCPMC7366071.

46. Chahraoui K, Bioy A, Cras E, Gilles F, Laurent A, Valache B, et al. [Psychological experience of health care professionals in intensive care unit: a qualitative and exploratory study]. Annales francaises d’anesthesie et de reanimation. 2011;30(4):342–8. Epub 2011/03/18. doi: 10.1016/j.annfar.2011.01.020. PubMed PMID: 21411265.

47. Krishnamoorthy Y, Nagarajan R, Saya GK, Menon V. Prevalence of psychological morbidities among general population, healthcare workers and COVID-19 patients amidst the COVID-19 pandemic: A systematic review and meta-analysis. Psychiatry research. 2020;293:113382. Epub 2020/08/24. doi: 10.1016/j.psychres.2020.113382. PubMed PMID: 32829073; PubMed Central PMCID: PMCPMC7417292.

48. Borges Migliavaca C, Stein C, Colpani V, Barker TH, Munn Z, Falavigna M. How are systematic reviews of prevalence conducted? A methodological study. BMC medical research methodology. 2020;20(1):96. Epub 2020/04/28. doi: 10.1186/s12874-020-00975-3. PubMed PMID: 32336279; PubMed Central PMCID: PMCPMC7184711.

49. Brudey C, Park J, Wiaderkiewicz J, Kobayashi I, Mellman TA, Marvar PJ. Autonomic and inflammatory consequences of posttraumatic stress disorder and the link to cardiovascular disease. American journal of physiology Regulatory, integrative and comparative physiology. 2015;309(4):R315–21. Epub 2015/06/13. doi: 10.1152/ajpregu.00343.2014. PubMed PMID: 26062635; PubMed Central PMCID: PMCPMC4538229.

50. Vandepitte S, Alleman T, Nopens I, Baetens J, Coenen S, De Smedt D. Cost-Effectiveness of COVID-19 Policy Measures: A Systematic Review. Value in health : the journal of the International Society for Pharmacoeconomics and Outcomes Research. 2021;24(11):1551–69. Epub 2021/10/30. doi: 10.1016/j.jval.2021.05.013. PubMed PMID: 34711355; PubMed Central PMCID: PMCPMC8481648.

51. Wen W, Chen C, Tang J, Wang C, Zhou M, Cheng Y, et al. Efficacy and safety of three new oral antiviral treatment (molnupiravir, fluvoxamine and Paxlovid) for COVID-19: a meta-analysis. Annals of medicine. 2022;54(1):516–23. Epub 2022/02/05. doi: 10.1080/07853890.2022.2034936. PubMed PMID: 35118917; PubMed Central PMCID: PMCPMC8820829.

52. Angeletti S, Giovanetti M, Fogolari M, Cella E, De Florio L, Lintas C, et al. SARS-CoV-2 AY.4.2 variant circulating in Italy: Genomic preliminary insight. Journal of medical virology. 2022;94(4):1689–92. Epub 2021/11/13. doi: 10.1002/jmv.27451. PubMed PMID: 34766651; PubMed Central PMCID: PMCPMC8661725.

53. Parums DV. Editorial: The XBB.1.5 (’Kraken’) Subvariant of Omicron SARS-CoV-2 and its Rapid Global Spread. Medical science monitor : international medical journal of experimental and clinical research. 2023;29:e939580. Epub 2023/02/02. doi: 10.12659/msm.939580. PubMed PMID: 36722047; PubMed Central PMCID: PMCPMC9901170.

54. Chen Q, Allot A, Lu Z. Keep up with the latest coronavirus research. Nature. 2020;579(7798):193. Epub 2020/03/12. doi: 10.1038/d41586-020-00694-1. PubMed PMID: 32157233.

55. Jung RG, Di Santo P, Clifford C, Prosperi-Porta G, Skanes S, Hung A, et al. Methodological quality of COVID-19 clinical research. Nat Commun. 2021;12(1):943. Epub 2021/02/13. doi: 10.1038/s41467-021-21220-5. PubMed PMID: 33574258; PubMed Central PMCID: PMCPMC7878793 Scientific and Edwards Lifesciences outside of the submitted work. The remaining authors declare no competing interests.

56. Burns PB, Rohrich RJ, Chung KC. The levels of evidence and their role in evidence-based medicine. Plast Reconstr Surg. 2011;128(1):305–10. Epub 2011/06/28. doi: 10.1097/PRS.0b013e318219c171. PubMed PMID: 21701348; PubMed Central PMCID: PMCPMC3124652.

57. Abu-Elenin MM. Immediate psychological outcomes associated with COVID-19 pandemic in frontline physicians: a cross-sectional study in Egypt. BMC psychiatry. 2021;21(1). doi: 10.1186/s12888-021-03225-y.

58. AlAteeq DA, Aljhani S, Althiyabi I, Majzoub S. Mental health among healthcare providers during coronavirus disease (COVID-19) outbreak in Saudi Arabia. Journal of infection and public health. 2020;13(10):1432–7. Epub 2020/09/17. doi: 10.1016/j.jiph.2020.08.013. PubMed PMID: 32933881; PubMed Central PMCID: PMCPMC7834809.

59. Ali S, Maguire S, Marks E, Doyle M, Sheehy C. Psychological impact of the COVID-19 pandemic on healthcare workers at acute hospital settings in the South-East of Ireland: an observational cohort multicentre study. BMJ open. 2020;10(12):e042930. Epub 2020/12/30. doi: 10.1136/bmjopen-2020-042930. PubMed PMID: 33371046; PubMed Central PMCID: PMCPMC7750872.

60. AlKandari S, Salman A, Al-Ghadban F, Ahmad R. A Cross-Sectional Study to Examine the Psychological Impact of the COVID-19 Pandemic on Healthcare Workers in Kuwait. International journal of environmental research and public health. 2022;19(17). doi: 10.3390/ijerph191710464.

61. Alkhamees AA, Assiri H, Alharbi HY, Nasser A, Alkhamees MA. Burnout and depression among psychiatry residents during COVID-19 pandemic. Human resources for health. 2021;19(1):46. Epub 2021/04/08. doi: 10.1186/s12960-021-00584-1. PubMed PMID: 33823857; PubMed Central PMCID: PMCPMC8022305.

62. Almalki AH, Alzahrani MS, Alshehri FS, Alharbi A, Alkhudaydi SF, Alshahrani RS, et al. The Psychological Impact of COVID-19 on Healthcare Workers in Saudi Arabia: A Year Later Into the Pandemic. Frontiers in Psychiatry. 2021;12. doi: 10.3389/fpsyt.2021.797545.

63. Alzahrani NS, Almarwani AM, Asiri SA, Alharbi HF, Alhowaymel FM. Factors influencing hospital anxiety and depression among emergency department nurses during the COVID-19 pandemic: A multi-center cross-sectional study. Frontiers in Psychiatry. 2022;13. doi: 10.3389/fpsyt.2022.912157.

64. Andlib S, Inayat S, Azhar K, Aziz F. Burnout and psychological distress among Pakistani nurses providing care to COVID-19 patients: A cross-sectional study. International nursing review. 2022. doi: 10.1111/inr.12750.

65. Arshad AR, Islam F. COVID-19 and Anxiety amongst Doctors: A Pakistani Perspective. Journal of the College of Physicians and Surgeons Pakistan. 2020;30(2):S106–S9. doi: 10.29271/JCPSP.2020.SUPP2.106.

66. Ashoor MM, Almulhem NJ, AlMubarak ZA, Alrahim AA, Alshammari SM, Alzahrani FS, et al. The psychological impact of the COVID-19 pandemic on otolaryngologists: Should we be concerned? Laryngoscope Investigative Otolaryngology. 2021;6(3):576–85. doi: 10.1002/lio2.556.

67. Awano N, Oyama N, Akiyama K, Inomata M, Kuse N, Tone M, et al. Anxiety, Depression, and Resilience of Healthcare Workers in Japan During the Coronavirus Disease 2019 Outbreak. Internal medicine (Tokyo, Japan). 2020;59(21):2693–9. Epub 2020/11/03. doi: 10.2169/internalmedicine.5694-20. PubMed PMID: 33132305; PubMed Central PMCID: PMCPMC7691033.

68. Azizi M, Kamali M, Moosazadeh M, Aarabi M, Ghasemian R, Hasannezhad Reskati M, et al. Assessing mental health status among Iranian healthcare workers in times of the COVID-19 pandemic: A web-based cross-sectional study. Brain and behavior. 2021;11(8). doi: 10.1002/brb3.2304.

69. Azoulay E, Pochard F, Reignier J, Argaud L, Bruneel F, Courbon P, et al. Symptoms of Mental Health Disorders in Critical Care Physicians Facing the Second COVID-19 Wave: A Cross-Sectional Study. Chest. 2021;160(3):944–55. doi: 10.1016/j.chest.2021.05.023. PubMed PMID: MEDLINE:34023323.

70. Babamiri M, Bashirian S, Khazaei S, Sohrabi MS, Heidarimoghadam R, Mortezapoor A, et al. Burnout and Mental Health of COVID-19 Frontline Healthcare Workers: Results from an Online Survey. Iranian Journal of Psychiatry. 2022;17(2):136–43. doi: 10.18502/ijps.v17i2.8903.

71. Baminiwatta A, De Silva S, Hapangama A, Basnayake K, Abayaweera C, Kulasinghe D, et al. Impact of COVID-19 on the mental health of frontline and non-frontline healthcare workers in Sri Lanka. The Ceylon medical journal. 2021;66(1):16–31. Epub 2022/01/06. doi: 10.4038/cmj.v66i1.9351. PubMed PMID: 34983177.

72. Cahill AG, Olshavsky ME, Newport DJ, Benzer J, Chambers KM, Custer J, et al. Occupational Risk Factors and Mental Health Among Frontline Health Care Workers in a Large US Metropolitan Area During the COVID-19 Pandemic. The primary care companion for CNS disorders. 2022;24(2). Epub 2022/03/12. doi: 10.4088/PCC.21m03166. PubMed PMID: 35276759.

73. Çalişkan F, Dost B. The evaluation of knowledge, attitudes, depression and anxiety levels among emergency physicians during the COVID-19 pandemic. Signa Vitae. 2020;16(1):163–71. doi: 10.22514/sv.2020.16.0022.

74. Carmassi C, Dell’Oste V, Barberi FM, Bertelloni CA, Pedrinelli V, Dell’Osso L. Mental Health Symptoms among General Practitioners Facing the Acute Phase of the COVID-19 Pandemic: Detecting Different Reaction Groups. International journal of environmental research and public health. 2022;19(7). Epub 2022/04/13. doi: 10.3390/ijerph19074007. PubMed PMID: 35409690; PubMed Central PMCID: PMCPMC8998411.

75. Carriero MC, Conte L, Calignano M, Lupo R, Calabrò A, Santoro P, et al. The psychological impact of the Coronavirus emergency on physicians and nurses: An Italian observational study. Acta Biomedica. 2021;92. doi: 10.23750/ABM.V92IS2.11575.

76. Chatterjee SS, Bhattacharyya R, Bhattacharyya S, Gupta S, Das S, Banerjee BB. Attitude, practice, behavior, and mental health impact of COVID-19 on doctors. Indian Journal of Psychiatry. 2020;62(3):257–65. doi: 10.4103/psychiatry.IndianJPsychiatry_333_20.

77. Chatzittofis A, Karanikola M, Michailidou K, Constantinidou A. Impact of the COVID-19 Pandemic on the Mental Health of Healthcare Workers. International journal of environmental research and public health. 2021;18(4). Epub 2021/02/07. doi: 10.3390/ijerph18041435. PubMed PMID: 33546513; PubMed Central PMCID: PMCPMC7913751.

78. Chen J, Liu X, Wang D, Jin Y, He M, Ma Y, et al. Risk factors for depression and anxiety in healthcare workers deployed during the COVID-19 outbreak in China. Social psychiatry and psychiatric epidemiology. 2021;56(1):47–55. doi: 10.1007/s00127-020-01954-1.

79. Chen L, Lin D, Feng H. An Investigation of Mental Health Status Among Medical Staff Following COVID-19 Outbreaks: A Cross-Sectional Study. Medical science monitor : international medical journal of experimental and clinical research. 2021;27:e929454. Epub 2021/07/01. doi: 10.12659/msm.929454. PubMed PMID: 34188013; PubMed Central PMCID: PMCPMC8256688.

80. Chen X, Arber A, Gao J, Zhang L, Ji M, Wang D, et al. The mental health status among nurses from low-risk areas under normalized COVID-19 pandemic prevention and control in China: A cross-sectional study. International journal of mental health nursing. 2021;30(4):975–87. doi: 10.1111/inm.12852. PubMed PMID: 151433613. Language: English. Entry Date: 20210720. Revision Date: 20220801. Publication Type: Article.

81. Chinvararak C, Kerdcharoen N, Pruttithavorn W, Polruamngern N, Asawaroekwisoot T, Munsukpol W, et al. Mental health among healthcare workers during COVID-19 pandemic in Thailand. PloS one. 2022;17(5):e0268704. Epub 2022/05/21. doi: 10.1371/journal.pone.0268704. PubMed PMID: 35594261; PubMed Central PMCID: PMCPMC9122199.

82. Civantos AM, Bertelli A, Gonçalves A, Getzen E, Chang C, Long Q, et al. Mental health among head and neck surgeons in Brazil during the COVID-19 pandemic: A national study. American Journal of Otolaryngology - Head and Neck Medicine and Surgery. 2020;41(6). doi: 10.1016/j.amjoto.2020.102694.

83. Çölkesen F, Çölkesen F. The effects of COVID-19 pandemic on the mental health of healthcare workers and recommendations for preventing loss of work efficiency. Erciyes Medical Journal. 2021;43(6):560–5. doi: 10.14744/etd.2021.03453.

84. Czepiel D, Hoek HW, van der Markt A, Rutten BPF, Veling W, Schirmbeck F, et al. The Association Between Exposure to COVID-19 and Mental Health Outcomes Among Healthcare Workers. Frontiers in public health. 2022;10:896843. Epub 2022/06/28. doi: 10.3389/fpubh.2022.896843. PubMed PMID: 35757645; PubMed Central PMCID: PMCPMC9226479.

85. d’Ussel M, Fels A, Durand X, Lemogne C, Chatellier G, Castreau N, et al. Factors associated with psychological symptoms in hospital workers of a French hospital during the COVID-19 pandemic: Lessons from the first wave. PloS one. 2022;17(4 April). doi: 10.1371/journal.pone.0267032.

86. Da’seh A, Obaid O, Rababa M. Psychological impact of coronavirus disease on nurses exposed and non-exposed to disease. International Journal of Africa Nursing Sciences. 2022;17. doi: 10.1016/j.ijans.2022.100442.

87. Dobson H, Malpas CB, Burrell AJC, Gurvich C, Chen L, Kulkarni J, et al. Burnout and psychological distress amongst Australian healthcare workers during the COVID-19 pandemic. Australasian Psychiatry. 2021;29(1):26–30. doi: 10.1177/1039856220965045.

88. Etesam F, Bafrani MA, Akbarpour S, Tarighatnia H, Rajabi G, Dolatshahi M, et al. Depression, Anxiety and Coping Responses among Iranian Healthcare Professionals during the Coronavirus Disease (COVID-19) Outbreak. Iranian Journal of Psychiatry. 2022;17(4):446–54. doi: 10.18502/ijps.v17i4.10694.

89. Frajerman A, Colle R, Hozer F, Deflesselle E, Rotenberg S, Chappell K, et al. Psychological distress among outpatient physicians in private practice linked to COVID-19 and related mental health during the second lockdown. Journal of psychiatric research. 2022;151:50–6. doi: 10.1016/j.jpsychires.2022.04.003. PubMed PMID: MEDLINE:35447507.

90. Fu M, Han D, Xu M, Mao C, Wang D. The psychological impact of anxiety and depression on Chinese medical staff during the outbreak of the COVID-19 pandemic: a cross-sectional study. Annals of palliative medicine. 2021;10(7):7759–74. doi: 10.21037/apm-21-1261.

91. GebreEyesus FA, Tarekegn TT, Amlak BT, Shiferaw BZ, Emeria MS, Geleta OT, et al. Levels and predictors of anxiety, depression, and stress during COVID-19 pandemic among frontline healthcare providers in Gurage zonal public hospitals, Southwest Ethiopia, 2020: A multicenter cross-sectional study. PloS one. 2021;16(11 November). doi: 10.1371/journal.pone.0259906.

92. Gilleen J, Santaolalla A, Valdearenas L, Salice C, Fusté M. Impact of the COVID-19 pandemic on the mental health and well-being of UK healthcare workers. BJPsych Open. 2021;7(3). doi: 10.1192/bjo.2021.42.

93. Goh ET, Denning M, Purkayastha S, Kinross J. Determinants of psychological well-being in healthcare workers during the covid-19 pandemic: A multinational cross-sectional study. British Journal of Surgery. 2021;108(SUPPL 2):ii23. doi: 10.1093/bjs/znab134.043.

94. Gonzalo RM, Ana RG, Patricia CA, Laura AL, Nathalia GT, Luis C, et al. Short-term emotional impact of COVID-19 pandemic on Spaniard health workers. Journal of affective disorders. 2021;278:390–4. doi: 10.1016/j.jad.2020.09.079.

95. Greenberg N, Weston D, Hall C, Caulfield T, Williamson V, Fong K. Mental health of staff working in intensive care during Covid-19. Occupational medicine (Oxford, England). 2021;71(2):62–7. Epub 2021/01/13. doi: 10.1093/occmed/kqaa220. PubMed PMID: 33434920; PubMed Central PMCID: PMCPMC7928568.

96. Guo WP, Min Q, Gu WW, Yu L, Xiao X, Yi WB, et al. Prevalence of mental health problems in frontline healthcare workers after the first outbreak of COVID-19 in China: a cross-sectional study. Health and Quality of Life Outcomes. 2021;19(1). doi: 10.1186/s12955-021-01743-7.

97. Habtamu Y, Admasu K, Tullu M, Damene W, Birhanu A, Beyero T, et al. Mental Health Outcomes among Frontline Health-Care Workers at Eka Kotebe National COVID-19 Treatment Center, Addis Ababa, Ethiopia, 2020: A Cross-Sectional Study. Neuropsychiatric Disease and Treatment. 2021;17:2831–40. doi: 10.2147/NDT.S311949.

98. Hayat K, Arshed M, Fiaz I, Afreen U, Khan FU, Khan TA, et al. Impact of COVID-19 on the Mental Health of Healthcare Workers: A Cross-Sectional Study From Pakistan. Frontiers in public health. 2021;9:603602. Epub 2021/05/14. doi: 10.3389/fpubh.2021.603602. PubMed PMID: 33981657; PubMed Central PMCID: PMCPMC8107369.

99. He L, Wang J, Zhang L, Wang F, Dong W, Zhao W. Risk Factors for Anxiety and Depressive Symptoms in Doctors During the Coronavirus Disease 2019 Pandemic. Frontiers in Psychiatry. 2021;12. doi: 10.3389/fpsyt.2021.687440.

100. Heesakkers H, Zegers M, van Mol MMC, van den Boogaard M. Mental well-being of intensive care unit nurses after the second surge of the COVID-19 pandemic: A cross-sectional and longitudinal study. Intensive & critical care nursing. 2022:103313. doi: 10.1016/j.iccn.2022.103313.

101. Hickling MT, Barnett SD. Psychological impact of COVID-19 on nursing personnel: A regional online survey. Journal of advanced nursing. 2022;78(9):3025–33. doi: 10.1111/jan.15339. PubMed PMID: WOS:000819350400001.

102. Hu D, Kong Y, Li W, Han Q, Zhang X, Zhu LX, et al. Frontline nurses’ burnout, anxiety, depression, and fear statuses and their associated factors during the COVID-19 outbreak in Wuhan, China: A large-scale cross-sectional study. EClinicalMedicine. 2020;24. doi: 10.1016/j.eclinm.2020.100424.

103. Jakhar J, Biswas PS, Kapoor M, Panghal A, Meena A, Fani H, et al. Comparative study of the mental health impact of the COVID-19 pandemic on health care professionals in India. Future microbiology. 2021;16:1267–76. Epub 2021/10/23. doi: 10.2217/fmb-2021-0084. PubMed PMID: 34674541; PubMed Central PMCID: PMCPMC8544479.

104. Jo SH, Koo BH, Seo WS, Yun SH, Kim HG. The psychological impact of the coronavirus disease pandemic on hospital workers in Daegu, South Korea. Comprehensive psychiatry. 2020;103:6. doi: 10.1016/j.comppsych.2020.152213. PubMed PMID: WOS:000594225900008.

105. Kafle B, Bagale Y, Kafle S, Parajuli A, Pandey S. Depression, anxiety and stress among healthcare workers during covid-19 pandemic in a tertiary care centre of nepal: A descriptive cross-sectional study. Journal of the Nepal Medical Association. 2021;59(235):239–42. doi: 10.31729/jnma.6083.

106. Kamali M, Azizi M, Moosazadeh M, Mehravaran H, Ghasemian R, Reskati MH, et al. Occupational burnout in Iranian health care workers during the COVID-19 pandemic. BMC psychiatry. 2022;22(1). doi: 10.1186/s12888-022-04014-x.

107. Kandouci C, Mecabih F, Mecabih I, Kadari C, Megherbi N, Achouri MY, et al. Psychosocial impact of COVID-19 among health workers in Algeria. Tunisie Medicale. 2021;99(11):1015–29.

108. Kapetanos K, Mazeri S, Constantinou D, Vavlitou A, Karaiskakis M, Kourouzidou D, et al. Exploring the factors associated with the mental health of frontline healthcare workers during the COVID-19 pandemic in Cyprus. PloS one. 2021;16(10):e0258475. Epub 2021/10/15. doi: 10.1371/journal.pone.0258475. PubMed PMID: 34648565; PubMed Central PMCID: PMCPMC8516220.

109. Khan N, Palepu A, Dodek P, Salmon A, Leitch H, Ruzycki S, et al. Cross-sectional survey on physician burnout during the COVID-19 pandemic in Vancouver, Canada: The role of gender, ethnicity and sexual orientation. BMJ open. 2021;11(5). doi: 10.1136/bmjopen-2021-050380.

110. Khatun MF, Parvin MF, Rashid MM, Alam MS, Kamrunnahar M, Talukder A, et al. Mental Health of Physicians During COVID-19 Outbreak in Bangladesh: A Web-Based Cross-Sectional Survey. Frontiers in public health. 2021;9:592058. Epub 2021/02/27. doi: 10.3389/fpubh.2021.592058. PubMed PMID: 33634065; PubMed Central PMCID: PMCPMC7902057.

111. Kiliç A, Gürcan MB, Aktura B, Şahin AR, Kökrek Z. Prevalence of anxiety and relationship of anxiety with coping styles and related factors in healthcare workers during COVID-19 pandemic. Psychiatria Danubina. 2021;33:161–71.

112. Lai J, Ma S, Wang Y, Cai Z, Hu J, Wei N, et al. Factors Associated With Mental Health Outcomes Among Health Care Workers Exposed to Coronavirus Disease 2019. JAMA network open. 2020;3(3):e203976. Epub 2020/03/24. doi: 10.1001/jamanetworkopen.2020.3976. PubMed PMID: 32202646; PubMed Central PMCID: PMCPMC7090843.

113. Lamiani G, Borghi L, Poli S, Razzini K, Colosio C, Vegni E. Hospital employees’ well-being six months after the covid-19 outbreak: Results from a psychological screening program in italy. International journal of environmental research and public health. 2021;18(11). doi: 10.3390/ijerph18115649.

114. Lasalvia A, Bodini L, Amaddeo F, Porru S, Carta A, Poli R, et al. The sustained psychological impact of the covid-19 pandemic on health care workers one year after the outbreak—a repeated cross-sectional survey in a tertiary hospital of north-east Italy. International journal of environmental research and public health. 2021;18(24). doi: 10.3390/ijerph182413374.

115. Li G, Miao J, Wang H, Xu S, Sun W, Fan Y, et al. Psychological impact on women health workers involved in COVID-19 outbreak in Wuhan: A cross-sectional study. Journal of Neurology, Neurosurgery and Psychiatry. 2020;91(8):895–7. doi: 10.1136/jnnp-2020-323134.

116. Li J, Guo J, Zhao J, Guo Y, Chen C. Influencing Factors of Mental Health Status of Dentists Under COVID-19 Epidemic. Frontiers in Psychiatry. 2022;13. doi: 10.3389/fpsyt.2022.933514.

117. Li S, Chai R, Wang Y, Wang J, Dong X, Xu H, et al. A survey of mental health status of obstetric nurses during the novel coronavirus pneumonia pandemic. Medicine. 2021;100(52):e28070. Epub 2021/12/31. doi: 10.1097/md.0000000000028070. PubMed PMID: 34967352; PubMed Central PMCID: PMCPMC8718244.

118. Li W, Mao X, Li J, Fang L, Du G, Qiao J, et al. Anxiety and Depression Among Imaging Doctors in Post-COVID-19 Period. SN Comprehensive Clinical Medicine. 2020;2(12):2595–9. doi: 10.1007/s42399-020-00654-w.

119. Li Y, Fan R, Lu Y, Li H, Liu X, Kong G, et al. Prevalence of psychological symptoms and associated risk factors among nurses in 30 provinces during the COVID-19 pandemic in China. The Lancet Regional Health - Western Pacific. 2022:100618. doi: https://doi.org/10.1016/j.lanwpc.2022.100618.

120. Liu CY, Yang YZ, Zhang XM, Xu X, Dou QL, Zhang WW, et al. The prevalence and influencing factors in anxiety in medical workers fighting COVID-19 in China: a cross-sectional survey. Epidemiology and infection. 2020;148:e98. Epub 2020/05/21. doi: 10.1017/s0950268820001107. PubMed PMID: 32430088; PubMed Central PMCID: PMCPMC7251286.

121. Lixia W, Xiaoming X, Lei S, Su H, Wo W, Xin F, et al. A cross-sectional study of the psychological status of 33,706 hospital workers at the late stage of the COVID-19 outbreak. Journal of affective disorders. 2022;297:156–68. doi: 10.1016/j.jad.2021.10.013.

122. Luceño-Moreno L, Talavera-Velasco B, García-Albuerne Y, Martín-García J. Symptoms of posttraumatic stress, anxiety, depression, levels of resilience and burnout in spanish health personnel during the COVID-19 pandemic. International journal of environmental research and public health. 2020;17(15):1–29. doi: 10.3390/ijerph17155514.

123. Martin J, Padierna A, Villanueva A, Quintana JM. Evaluation of the mental health of health professionals in the COVID-19 era. What mental health conditions are our health care workers facing in the new wave of coronavirus? International journal of clinical practice. 2021;75(10):e14607. doi: 10.1111/ijcp.14607. PubMed PMID: MEDLINE:34231287.

124. Martínez Pajuelo AR, Irrazabal Ramos JE, Lazo-Porras M. Anxiety, Depression, and Post-Traumatic Stress Disorder (PTSD) Symptomatology According to Gender in Health-Care Workers during the COVID-19 Pandemic in Peru Shortened Title: “Psychological Impact of the Pandemic on Women”. International journal of environmental research and public health. 2022;19(19). doi: 10.3390/ijerph191911957.

125. Martínez-Caballero CM, Cárdaba-García RM, Varas-Manovel R, García-Sanz LM, Martínez-Piedra J, Fernández-Carbajo JJ, et al. Analyzing the impact of COVID-19 trauma on developing post-traumatic stress disorder among emergency medical workers in Spain. International journal of environmental research and public health. 2021;18(17). doi: 10.3390/ijerph18179132.

126. Martsenkovskyi D, Babych V, Martsenkovska I, Napryeyenko O, Napryeyenko N, Martsenkovsky I. Depression, anxiety, stress and trauma-related symptoms and their association with perceived social support in medical professionals during the COVID-19 pandemic in Ukraine. Postepy Psychiatrii i Neurologii. 2022;31(1):6–14. doi: 10.5114/ppn.2022.114657.

127. Mattila E, Peltokoski J, Neva MH, Kaunonen M, Helminen M, Parkkila A-K. COVID-19: anxiety among hospital staff and associated factors. Annals of medicine. 2021;53(1):237–46. doi: 10.1080/07853890.2020.1862905. PubMed PMID: MEDLINE:33350869.

128. McGuinness SL, Johnson J, Eades O, Cameron PA, Forbes A, Fisher J, et al. Mental Health Outcomes in Australian Healthcare and Aged-Care Workers during the Second Year of the COVID-19 Pandemic. International journal of environmental research and public health. 2022;19(9). Epub 2022/05/15. doi: 10.3390/ijerph19094951. PubMed PMID: 35564351; PubMed Central PMCID: PMCPMC9103405.

129. Mekhemar M, Attia S, Dörfer C, Conrad J. Dental Nurses’ Mental Health in Germany: A Nationwide Survey during the COVID-19 Pandemic. International journal of environmental research and public health. 2021;18(15). Epub 2021/08/08. doi: 10.3390/ijerph18158108. PubMed PMID: 34360401; PubMed Central PMCID: PMCPMC8345776.

130. Mekhemar M, Attia S, Dörfer C, Conrad J. The psychological impact of the covid-19 pandemic on dentists in germany. Journal of Clinical Medicine. 2021;10(5):1–18. doi: 10.3390/jcm10051008.

131. Mennicken B, Petit G, Yombi J-C, Belkhir L, Deschietere G, Germeau N, et al. Psychological distress among hospital caregivers during and after the first wave of COVID-19: Individual factors involved in the severity of symptoms expression. Psychiatry Research Communications. 2022;2(2):100037. doi: https://doi.org/10.1016/j.psycom.2022.100037.

132. Meo SA, Alkhalifah JM, Alshammari NF, Alnufaie WS. Comparison of generalized anxiety and sleep disturbance among frontline and second-line healthcare workers during the covid-19 pandemic. International journal of environmental research and public health. 2021;18(11). doi: 10.3390/ijerph18115727.

133. Mi T, Yang X, Sun S, Li X, Tam CC, Zhou Y, et al. Mental Health Problems of HIV Healthcare Providers During the COVID-19 Pandemic: The Interactive Effects of Stressors and Coping. AIDS and behavior. 2021;25(1):18–27. Epub 2020/11/01. doi: 10.1007/s10461-020-03073-z. PubMed PMID: 33128108; PubMed Central PMCID: PMCPMC7598225.

134. Morawa E, Schug C, Geiser F, Beschoner P, Jerg-Bretzke L, Albus C, et al. Psychosocial burden and working conditions during the COVID-19 pandemic in Germany: The VOICE survey among 3678 health care workers in hospitals. Journal of psychosomatic research. 2021;144:110415. Epub 2021/03/21. doi: 10.1016/j.jpsychores.2021.110415. PubMed PMID: 33743398; PubMed Central PMCID: PMCPMC7944879.

135. Moro MF, Calamandrei G, Poli R, Di Mattei V, Perra A, Kurotschka PK, et al. The Impact of the COVID-19 Pandemic on the Mental Health of Healthcare Workers in Italy: Analyzing the Role of Individual and Workplace-Level Factors in the Reopening Phase After Lockdown. Frontiers in Psychiatry. 2022;13. doi: 10.3389/fpsyt.2022.867080.

136. Mosheva M, Gross R, Hertz-Palmor N, Hasson-Ohayon I, Kaplan R, Cleper R, et al. The association between witnessing patient death and mental health outcomes in frontline COVID-19 healthcare workers. Depression and anxiety. 2021;38(4):468–79. doi: 10.1002/da.23140.

137. Motahedi S, Aghdam NF, Khajeh M, Baha R, Aliyari R, Bagheri H, et al. Anxiety and depression among healthcare workers during COVID-19 pandemic: A cross-sectional study. Heliyon. 2021;7(12):e08570. doi: https://doi.org/10.1016/j.heliyon.2021.e08570.

138. Mulatu HA, Tesfaye M, Woldeyes E, Bayisa T, Fisseha H, Kassu RA. The prevalence of common mental disorders among healthcare professionals during the COVID-19 pandemic at a tertiary Hospital in Addis Ababa, Ethiopia. Journal of Affective Disorders Reports. 2021;6:100246. doi: https://doi.org/10.1016/j.jadr.2021.100246.

139. Nagarkar R, Patil R, Gadade K, Paleja N, Ramesh YV. Psychological and Mental Health Burden on Health Care Providers in a Cancer Centre during COVID-19 Pandemic Outbreak in India. Psychiatria Danubina. 2022;34(1):164–70. Epub 2022/04/26. doi: 10.24869/psyd.2022.164. PubMed PMID: 35467634.

140. Nguyen TT, Le XTT, Nguyen NTT, Nguyen QN, Le HT, Pham QT, et al. Psychosocial Impacts of COVID-19 on Healthcare Workers During the Nationwide Partial Lockdown in Vietnam in April 2020. Frontiers in Psychiatry. 2021;12. doi: 10.3389/fpsyt.2021.562337.

141. Ning X, Yu F, Huang Q, Li X, Luo Y, Huang Q, et al. The mental health of neurological doctors and nurses in Hunan Province, China during the initial stages of the COVID-19 outbreak. BMC psychiatry. 2020;20(1). doi: 10.1186/s12888-020-02838-z.

142. Nordin S, Yaacob NA, Kelak J, Ilyas AH, Daud A. The Mental Health of Malaysia’s Northwest Healthcare Workers during the Relaxation of COVID-19 Restrictions and Its Associated Factors. International journal of environmental research and public health. 2022;19(13). Epub 2022/07/10. doi: 10.3390/ijerph19137794. PubMed PMID: 35805454; PubMed Central PMCID: PMCPMC9266069.

143. NS AL, AH AL, SS AL, Hersi AS. Depression among physicians and other medical employees involved in the COVID-19 outbreak: A cross-sectional study. Medicine. 2021;100(15):e25290. Epub 2021/04/14. doi: 10.1097/md.0000000000025290. PubMed PMID: 33847627; PubMed Central PMCID: PMCPMC8052024.

144. Olivares-Tirado P, Zanga-Pizarro R. Impact of COVID-19 pandemic outbreak on mental health of the hospital front-line healthcare workers in Chile: a difference-in-differences approach. Journal of public health (Oxford, England). 2022. doi: 10.1093/pubmed/fdac008.

145. Osório FL, Silveira ILM, Pereira-Lima K, Crippa JADS, Hallak JEC, Zuardi AW, et al. Risk and Protective Factors for the Mental Health of Brazilian Healthcare Workers in the Frontline of COVID-19 Pandemic. Frontiers in Psychiatry. 2021;12. doi: 10.3389/fpsyt.2021.662742.

146. Oteir AO, Nazzal MS, Jaber AF, Alwidyan MT, Raffee LA. Depression, anxiety and insomnia among frontline healthcare workers amid the coronavirus pandemic (COVID-19) in Jordan: a cross-sectional study. BMJ open. 2022;12(1):e050078. Epub 2022/02/03. doi: 10.1136/bmjopen-2021-050078. PubMed PMID: 35105616; PubMed Central PMCID: PMCPMC8804306.

147. Pan X, Xiao Y, Ren D, Xu ZM, Zhang Q, Yang LY, et al. Prevalence of mental health problems and associated risk factors among military healthcare workers in specialized COVID-19 hospitals in Wuhan, China: A cross-sectional survey. Asia-Pacific psychiatry : official journal of the Pacific Rim College of Psychiatrists. 2022;14(1):e12427. Epub 2020/10/23. doi: 10.1111/appy.12427. PubMed PMID: 33089622; PubMed Central PMCID: PMCPMC7645907.

148. Pandey A, Sharma C, Chapagain RH, Devkota N, Ranabhat K, Pant S, et al. Stress, Anxiety, Depression and Their Associated Factors among Health Care Workers During COVID -19 Pandemic in Nepal. Journal of Nepal Health Research Council. 2021;18(4):655–60. Epub 2021/01/30. doi: 10.33314/jnhrc.v18i4.3190. PubMed PMID: 33510505.

149. Paniagua-Ávila A, Ramírez DE, Barrera-Pérez A, Calgua E, Castro C, Peralta-García A, et al. Mental health of Guatemalan health care workers during the COVID-19 pandemic: baseline findings from the HEROES cohort study. Revista Panamericana de Salud Publica/Pan American Journal of Public Health. 2022;46. doi: 10.26633/RPSP.2022.79.

150. Pappa S, Athanasiou N, Sakkas N, Patrinos S, Sakka E, Barmparessou Z, et al. From recession to depression? Prevalence and correlates of depression, anxiety, traumatic stress and burnout in healthcare workers during the covid-19 pandemic in greece: a multi-center, cross-sectional study. International journal of environmental research and public health. 2021;18(5):1–16. doi: 10.3390/ijerph18052390.

151. Parthasarathy R, Ts J, K T, Murthy P. Mental health issues among health care workers during the COVID-19 pandemic – A study from India. Asian journal of psychiatry. 2021;58. doi: 10.1016/j.ajp.2021.102626.

152. Pelissier C, Paredes J, Moulin M, Bitot T, Fakra E, Fontana L. Telework and psychological health in hospital staff during the first wave of the COVID-19 epidemic in France. International journal of environmental research and public health. 2021;18(19). doi: 10.3390/ijerph181910433.

153. Pérez Herrera LC, Peñaranda D, Moreno-López S, Garcia E, Garcia IP, Peñaranda A. Mental health and daily and occupational activities of otolaryngologists in Colombia. Otolaryngology - Head and Neck Surgery. 2021;165(1 SUPPL):P323. doi: 10.1177/01945998211030910h.

154. Pisanu E, Di Benedetto A, Infurna MR, Rumiati RI. Psychological Impact in Healthcare Workers During Emergencies: The Italian Experience With COVID-19 First Wave. Frontiers in Psychiatry. 2022;13. doi: 10.3389/fpsyt.2022.818674.

155. Proserpio P, Zambrelli E, Lanza A, Dominese A, Di Giacomo R, Quintas R, et al. Sleep disorders and mental health in hospital workers during the COVID-19 pandemic: a cross-sectional multicenter study in Northern Italy. Neurological Sciences. 2022;43(4):2241–51. doi: 10.1007/s10072-021-05813-y.

156. Rahman A, Deeba F, Akhter S, Bashar F, Nomani D, Koot J, et al. Mental health condition of physicians working frontline with COVID-19 patients in Bangladesh. BMC psychiatry. 2021;21(1):615. Epub 2021/12/11. doi: 10.1186/s12888-021-03629-w. PubMed PMID: 34886844; PubMed Central PMCID: PMCPMC8655324.

157. Repon MAU, Pakhe SA, Quaiyum S, Das R, Daria S, Islam MR. Effect of COVID-19 pandemic on mental health among Bangladeshi healthcare professionals: A cross-sectional study. Science progress. 2021;104(2):368504211026409. Epub 2021/06/25. doi: 10.1177/00368504211026409. PubMed PMID: 34166132.

158. Riaz B, Rafai WA, Ussaid A, Masood A, Anwar S, Baig FA, et al. The psychological impact of COVID-19 on healthcare workers in Pakistan. Future Healthcare Journal. 2021;8(2):E293–E8. doi: 10.7861/FHJ.2020-0193.

159. RilleraMarzo R, Villanueva EQ, Chandra U, Htay MNN, Shrestha R, Shrestha S. Risk Perception, Mental Health Impacts and Coping Strategies during Covid-19 Pandemic among Filipino Healthcare Workers. Journal of Public Health Research. 2021;10(2_suppl). doi: 10.4081/jphr.2021.2604.

160. Rossi R, Socci V, Pacitti F, Di Lorenzo G, Di Marco A, Siracusano A, et al. Mental Health Outcomes Among Frontline and Second-Line Health Care Workers During the Coronavirus Disease 2019 (COVID-19) Pandemic in Italy. JAMA network open. 2020;3(5):e2010185–e. doi: 10.1001/jamanetworkopen.2020.10185. PubMed PMID: 143494368. Language: English. Entry Date: 20200602. Revision Date: 20200629. Publication Type: Article.

161. Sabbaghi M, Miri K, Kahi R, Nia MN. Investigation of stress, anxiety, and depression levels of Pre-Hospital Emergency Medicine personnel in eastern Iran during the Covid-19 pandemic. BMC emergency medicine. 2022;22(1). doi: 10.1186/s12873-022-00647-z.

162. Salehiniya H, Abbaszadeh H. Prevalence of corona-associated anxiety and mental health disorder among dentists during the COVID-19 pandemic. Neuropsychopharmacology Reports. 2021;41(2):223–9. doi: 10.1002/npr2.12179.

163. Salman M, Mustafa ZU, Raza MH, Khan TM, Asif N, Tahir H, et al. Psychological Effects of COVID-19 among Healthcare Workers and How They Are Coping: A Web-based, Cross-sectional Study during the First Wave of COVID-19 in Pakistan. Disaster medicine and public health preparedness. 2022:1–13. doi: 10.1017/dmp.2022.4.

164. Samir AlKudsi Z, Hany Kamel N, El-Awaisi A, Shraim M, Saffouh El Hajj M. Mental health, burnout and resilience in community pharmacists during the COVID-19 pandemic: A cross-sectional study. Saudi Pharmaceutical Journal. 2022;30(7):1009–17. doi: 10.1016/j.jsps.2022.04.015.

165. Sertoz OO, Tuncel OK, Sertoz N, Hepdurgun C, Haznedaroglu DI, Bor C. Burnout in Healthcare Professionals During the Covid-19 Pandemic in a Tertiary Care University Hospital: Evaluation of the Need for Psychological Support. Turk Psikiyatr Derg. 2021;32(2):75–86. doi: 10.5080/u25964. PubMed PMID: WOS:000697193200001.

166. Setiawati Y, Wahyuhadi J, Joestandari F, Maramis MM, Atika A. Anxiety and resilience of healthcare workers during COVID-19 pandemic in Indonesia. Journal of Multidisciplinary Healthcare. 2021;14:1–8. doi: 10.2147/JMDH.S276655.

167. Shah ED, Pourmorteza M, Elmunzer BJ, Ballou SK, Papachristou GI, Lara LF, et al. Psychological Health Among Gastroenterologists During the COVID-19 Pandemic: A National Survey. Clinical gastroenterology and hepatology : the official clinical practice journal of the American Gastroenterological Association. 2021;19(4):836–8.e3. Epub 2020/12/06. doi: 10.1016/j.cgh.2020.11.043. PubMed PMID: 33278574; PubMed Central PMCID: PMCPMC7955767.

168. Shah J, Monroe-Wise A, Talib Z, Nabiswa A, Said M, Abeid A, et al. Mental health disorders among healthcare workers during the COVID-19 pandemic: a cross-sectional survey from three major hospitals in Kenya. BMJ open. 2021;11(6):e050316. Epub 2021/06/11. doi: 10.1136/bmjopen-2021-050316. PubMed PMID: 34108174; PubMed Central PMCID: PMCPMC8190985.

169. Shah N, Raheem A, Sideris M, Velauthar L, Saeed F. Mental health amongst obstetrics and gynaecology doctors during the COVID-19 pandemic: Results of a UK-wide study. European Journal of Obstetrics and Gynecology and Reproductive Biology. 2020;253:90–4. doi: 10.1016/j.ejogrb.2020.07.060.

170. Shechter A, Chiuzan C, Shang Y, Ko G, Diaz F, Venner HK, et al. Prevalence, incidence, and factors associated with posttraumatic stress at three-month follow-up among new york city healthcare workers after the first wave of the COVID-19 pandemic. International journal of environmental research and public health. 2022;19(1). doi: 10.3390/ijerph19010262.

171. Shechter A, Diaz F, Moise N, Anstey DE, Ye S, Agarwal S, et al. Psychological distress, coping behaviors, and preferences for support among New York healthcare workers during the COVID-19 pandemic. General hospital psychiatry. 2020;66:1–8. doi: 10.1016/j.genhosppsych.2020.06.007.

172. Shen H, Wang H, Zhou F, Chen J, Deng L. Mental health status of medical staff in the epidemic period of coronavirus disease 2019. Zhong nan da xue xue bao Yi xue ban = Journal of Central South University Medical sciences. 2020;45(6):633–40. Epub 2020/09/04. doi: 10.11817/j.issn.1672-7347.2020.200070. PubMed PMID: 32879119.

173. Shrestha SL. Prevalence of Psychological Effect of COVID-19 on Medical Professionals in a Tertiary Care Center. JNMA; journal of the Nepal Medical Association. 2020;58(228):550–3. Epub 2020/09/25. doi: 10.31729/jnma.5087. PubMed PMID: 32968286; PubMed Central PMCID: PMCPMC7580372.

174. Siamisang K, Kebadiretse D, Tjirare LT, Muyela C, Gare K, Masupe T. Prevalence and predictors of depression, anxiety and stress among frontline healthcare workers at COVID-19 isolation sites in Gaborone, Botswana. PloS one. 2022;17(8):e0273052. Epub 2022/08/24. doi: 10.1371/journal.pone.0273052. PubMed PMID: 35998130; PubMed Central PMCID: PMCPMC9398030.

175. Simonetti V, Durante A, Ambrosca R, Arcadi P, Graziano G, Pucciarelli G, et al. Anxiety, sleep disorders and self-efficacy among nurses during COVID-19 pandemic: A large cross-sectional study. Journal of clinical nursing. 2021;30(9-10):1360–71. doi: 10.1111/jocn.15685. PubMed PMID: WOS:000617966300001.

176. Singh J, Sood M, Chadda RK, Singh V, Kattula D. Mental health issues and coping among health care workers during COVID19 pandemic: Indian perspective. Asian journal of psychiatry. 2021;61:102685. doi: https://doi.org/10.1016/j.ajp.2021.102685.

177. Sitanggang FP, Wirawan GBS, Wirawan IMA, Lesmana CBJ, Januraga PP. Determinants of mental health and practice behaviors of general practitioners during covid-19 pandemic in Bali, Indonesia: A cross-sectional study. Risk Management and Healthcare Policy. 2021;14:2055–64. doi: 10.2147/RMHP.S305373.

178. Smallwood N, Karimi L, Bismark M, Putland M, Johnson D, Dharmage SC, et al. High levels of psychosocial distress among Australian frontline healthcare workers during the COVID-19 pandemic: A cross-sectional survey. General Psychiatry. 2021;34(5). doi: 10.1136/gpsych-2021-100577.

179. Sobregrau Sangra P, Aguilo Mir S, Castro Ribeiro T, Esteban-Sepulveda S, Garcia Pages E, Lopez Barbeito B, et al. Mental health assessment of Spanish healthcare workers during the SARS-CoV-2 pandemic. A cross-sectional study. Comprehensive psychiatry. 2022;112:152278. doi: 10.1016/j.comppsych.2021.152278. PubMed PMID: MEDLINE:34678607.

180. Song X, Fu W, Liu X, Luo Z, Wang R, Zhou N, et al. Mental health status of medical staff in emergency departments during the Coronavirus disease 2019 epidemic in China. Brain, behavior, and immunity. 2020;88:60–5. Epub 2020/06/09. doi: 10.1016/j.bbi.2020.06.002. PubMed PMID: 32512134; PubMed Central PMCID: PMCPMC7273140.

181. Srikanth P, Monsey LM, Meischke HW, Baker MG. Determinants of Stress, Depression, Quality of Life, and Intent to Leave in Washington State Emergency Medical Technicians During COVID-19. Journal of occupational and environmental medicine. 2022;64(8):642–8. doi: 10.1097/JOM.0000000000002587.

182. Stocchetti N, Segre G, Zanier ER, Zanetti M, Campi R, Scarpellini F, et al. Burnout in intensive care unit workers during the second wave of the covid-19 pandemic: A single center cross-sectional Italian study. International journal of environmental research and public health. 2021;18(11). doi: 10.3390/ijerph18116102.

183. Su PA, Lo MC, Wang CL, Yang PC, Chang CI, Huang MC, et al. The correlation between professional quality of life and mental health outcomes among hospital personnel during the Covid-19 pandemic in Taiwan. Journal of Multidisciplinary Healthcare. 2021;14:3485–95. doi: 10.2147/JMDH.S330533.

184. Su Q, Ma X, Liu S, Liu S, Goodman BA, Yu M, et al. Adverse Psychological Reactions and Psychological Aids for Medical Staff During the COVID-19 Outbreak in China. Frontiers in Psychiatry. 2021;12. doi: 10.3389/fpsyt.2021.580067.

185. Sun D, Yang D, Li Y, Zhou J, Wang W, Wang Q, et al. Psychological impact of 2019 novel coronavirus (2019-nCoV) outbreak in health workers in China. Epidemiology and infection. 2020;148:e96. Epub 2020/05/21. doi: 10.1017/s0950268820001090. PubMed PMID: 32430086; PubMed Central PMCID: PMCPMC7251284.

186. Suryavanshi N, Kadam A, Dhumal G, Nimkar S, Mave V, Gupta A, et al. Mental health and quality of life among healthcare professionals during the COVID-19 pandemic in India. Brain and behavior. 2020;10(11):e01837. Epub 2020/09/13. doi: 10.1002/brb3.1837. PubMed PMID: 32918403; PubMed Central PMCID: PMCPMC7667343.

187. Syamlan AT, Salamah S, Alkaff FF, Prayudi YE, Kamil M, Irzaldy A, et al. Mental health and health-related quality of life among healthcare workers in Indonesia during the COVID-19 pandemic: a cross-sectional study. BMJ open. 2022;12(4):e057963. Epub 2022/04/10. doi: 10.1136/bmjopen-2021-057963. PubMed PMID: 35396304; PubMed Central PMCID: PMCPMC8996007.

188. Maslach C, Jackson SE. The measurement of experienced burnout. Journal of Organizational Behavior. 1981;2(2):99–113. doi: 10.1002/job.4030020205.

189. Demerouti E, Bakker AB, Vardakou I, Kantas A. The convergent validity of two burnout instruments: A multitrait-multimethod analysis. European Journal of Psychological Assessment. 2003;19(1):12–23. doi: 10.1027/1015-5759.19.1.12.

190. Tan BYQ, Chew NWS, Lee GKH, Jing M, Goh Y, Yeo LLL, et al. Psychological impact of the COVID-19 pandemic on health care workers in Singapore. Annals of internal medicine. 2020;173(4):317–20. doi: 10.7326/M20-1083.

191. Tan YQ, Wang Z, Yap QV, Chan YH, Ho RC, Hamid ARAH, et al. Psychological Health of Surgeons in a Time of COVID-19: A Global Survey. Annals of surgery. 2021. doi: 10.1097/SLA.0000000000004775.

192. Tang J, Wu Y, Qi H, Li D, Shi J, Wang W, et al. Psychological outcomes and associated factors amongst healthcare workers during a single wave, deeper into the COVID-19 pandemic in China. Frontiers in Psychiatry. 2022;13. doi: 10.3389/fpsyt.2022.983909.

193. Tao J, Lin Y, Jiang L, Zhou Z, Zhao J, Qu D, et al. Psychological Impact of the COVID-19 Pandemic on Emergency Dental Care Providers on the Front Lines in China. International Dental Journal. 2021;71(3):197–205. doi: https://doi.org/10.1016/j.identj.2020.12.001.

194. Taşdelen R, Ayik B, Kaya H, Ercis M, Ertekin E. Psychological Reactions of Turkish Healthcare Workers During Covid-19 Outbreak: The Impact of Stigmatization. Noropsikiyatri Arsivi. 2022;59(2):133–8. doi: 10.29399/npa.27785.

195. Than HM, Nong VM, Nguyen CT, Dong KP, Ngo HT, Doan TT, et al. Mental health and health-related quality-of-life outcomes among frontline health workers during the peak of covid-19 outbreak in Vietnam: A cross-sectional study. Risk Management and Healthcare Policy. 2020;13:2927–36. doi: 10.2147/RMHP.S280749.

196. Turan O, Demirci NY, Güntülü AK, Akçay Ş, Aktürk ÜA, Bilaçeroğlu S, et al. Anxiety and depression levels of healthcare workers during Covid-19 pandemic. African health sciences. 2022;22(1):532–40. doi: 10.4314/ahs.v22i1.62.

197. Uyaroğlu OA, Başaran N, Ozisik L, Karahan S, Tanriover MD, Guven GS, et al. Evaluation of the effect of COVID-19 pandemic on anxiety severity of physicians working in the internal medicine department of a tertiary care hospital: a cross-sectional survey. Internal medicine journal. 2020;50(11):1350–8. Epub 2020/10/03. doi: 10.1111/imj.14981. PubMed PMID: 33006419; PubMed Central PMCID: PMCPMC7537014.

198. Uz B, Savaşan E, Soğancı D. Anxiety, Depression and Burnout Levels of Turkish Healthcare Workers at the End of the First Period of COVID-19 Pandemic in Turkey. Clinical Psychopharmacology and Neuroscience. 2022;20(1):97–108. doi: 10.9758/CPN.2022.20.1.97.

199. Vadi S, Shah S, Bajpe S, George N, Santhosh A, Sanwalka N, et al. Mental Health Indices of Intensive Care Unit and Emergency Room Frontliners during the Severe Acute Respiratory Syndrome Coronavirus 2 Pandemic in India. Indian Journal of Critical Care Medicine. 2022;26(1):100–7. doi: 10.5005/jp-journals-10071-24081.

200. Valaine L, Ancāne G, Utināns A, Briģis Ģ. Mental Health and Associated Demographic and Occupational Factors among Health Care Workers during the COVID-19 Pandemic in Latvia. Medicina (Kaunas, Lithuania). 2021;57(12). Epub 2021/12/25. doi: 10.3390/medicina57121381. PubMed PMID: 34946326; PubMed Central PMCID: PMCPMC8705324.

201. van de Venter R, Williams R, Stindt C, ten Ham-Baloyi W. Coronavirus-related anxiety and fear among South African diagnostic radiographers working in the clinical setting during the pandemic. Journal of Medical Imaging and Radiation Sciences. 2021;52(4):586–94. doi: 10.1016/j.jmir.2021.09.016.

202. Van Wert MJ, Gandhi S, Gupta I, Singh A, Eid SM, Haroon Burhanullah M, et al. Healthcare Worker Mental Health After the Initial Peak of the COVID-19 Pandemic: a US Medical Center Cross-Sectional Survey. Journal of general internal medicine. 2022;37(5):1169–76. Epub 2022/01/08. doi: 10.1007/s11606-021-07251-0. PubMed PMID: 34993856; PubMed Central PMCID: PMCPMC8734540.

203. Vancappel A, Jansen E, Ouhmad N, Desmidt T, Etain B, Bergey C, et al. Psychological Impact of Exposure to the COVID-19 Sanitary Crisis on French Healthcare Workers: Risk Factors and Coping Strategies. Frontiers in Psychiatry. 2021;12. doi: 10.3389/fpsyt.2021.701127.

204. Wanigasooriya K, Palimar P, Naumann DN, Ismail K, Fellows JL, Logan P, et al. Mental health symptoms in a cohort of hospital healthcare workers following the first peak of the COVID-19 pandemic in the UK. BJPsych Open. 2020;7(1). doi: 10.1192/bjo.2020.150.

205. Wayessa ZJ, Melesse GT, Amaje Hadona E, Wako WG. Prevalence of depressive symptoms due to COVID-19 and associated factors among healthcare workers in Southern Ethiopia. SAGE Open Medicine. 2021;9. doi: 10.1177/20503121211032810.

206. Weibelzahl S, Reiter J, Duden G. Depression and Anxiety in Healthcare Professionals during the COVID-19 Pandemic. Epidemiology and infection. 2021. doi: 10.1017/S0950268821000303.

207. Xiong H, Yi S, Lin Y. The Psychological Status and Self-Efficacy of Nurses During COVID-19 Outbreak: A Cross-Sectional Survey. Inquiry : a journal of medical care organization, provision and financing. 2020;57:46958020957114. Epub 2020/09/10. doi: 10.1177/0046958020957114. PubMed PMID: 32900271; PubMed Central PMCID: PMCPMC7485150.

208. Yadeta TA, Dessie Y, Balis B. Magnitude and Predictors of Health Care Workers Depression During the COVID-19 Pandemic: Health Facility-Based Study in Eastern Ethiopia. Frontiers in Psychiatry. 2021;12. doi: 10.3389/fpsyt.2021.654430.

209. Yang Y, Liu D, Liu B, Ou W, Wang L, Ma Y, et al. Prevalence of Post-traumatic Stress Disorder Status Among Healthcare Workers and Its Impact on Their Mental Health During the Crisis of COVID-19: A Cross-Sectional Study. Frontiers in public health. 2022;10:904550. Epub 2022/08/06. doi: 10.3389/fpubh.2022.904550. PubMed PMID: 35928490; PubMed Central PMCID: PMCPMC9343759.

210. Youssfi I, Mechergui N, Merchaoui I, Bouden F, Said HB, Youssef I, et al. Perception of mental health and professional quality of life in Tunisian doctors during the COVID-19 pandemic: a descriptive cross-sectional study. The Pan African medical journal. 2021;40:139. Epub 2021/12/16. doi: 10.11604/pamj.2021.40.139.30358. PubMed PMID: 34909107; PubMed Central PMCID: PMCPMC8641639.

211. Yu B, Barnett D, Menon V, Rabiee L, De Castro YS, Kasubhai M, et al. Healthcare worker trauma and related mental health outcomes during the COVID-19 outbreak in New York City. PloS one. 2022;17(4 April). doi: 10.1371/journal.pone.0267315.

212. Zarzour M, Hachem C, Kerbage H, Richa S, Choueifaty DE, Saliba G, et al. Anxiety and sleep quality in a sample of Lebanese healthcare workers during the COVID-19 outbreak. L’Encephale. 2022;48(5):496–503. doi: 10.1016/j.encep.2021.06.016.

213. Edmondson D, von Känel R. Post-traumatic stress disorder and cardiovascular disease. The Lancet Psychiatry. 2017;4(4):320–329. doi: 10.1016/S2215-0366(16)30377-7.

214. Zhan J, Chen C, Yan X, Wei X, Zhan L, Chen H, et al. Relationship between social support, anxiety, and depression among frontline healthcare workers in China during COVID-19 pandemic. Frontiers in Psychiatry. 2022;13. doi: 10.3389/fpsyt.2022.947945.

215. World Health Organization. WHO validates 11th vaccine for COVID-19. World Health Organization. 2022. Available from: https://www.who.int/news/item/2022-04-14-who-validates-11th-vaccine-for-covid-19

216. Zhang H, Shi Y, Jing P, Zhan P, Fang Y, Wang F. Posttraumatic stress disorder symptoms in healthcare workers after the peak of the COVID-19 outbreak: A survey of a large tertiary care hospital in Wuhan. Psychiatry research. 2020;294:113541. Epub 2020/11/01. doi: 10.1016/j.psychres.2020.113541. PubMed PMID: 33128999; PubMed Central PMCID: PMCPMC7585629.

217. World Health Organization. Statement on the fourteenth meeting of the International Health Regulations (2005) Emergency Committee regarding the coronavirus disease (COVID-19) pandemic. World Health Organization. 2023. Available from: https://www.who.int/news/item/2023/statement-on-the-fourteenth-meeting-of-the-international-health-regulations-(2005)-emergency-committee-regarding-the-coronavirus-disease-(covid-19)-pandemic

218. Zhang J, Song S, Zhang M, Wang R. Influencing factors for mental health of general practitioners in Hebei Province under the outbreak of COVID-19: A cross-sectional study. International journal of clinical practice. 2021;75(11):e14783. Epub 2021/09/06. doi: 10.1111/ijcp.14783. PubMed PMID: 34482597; PubMed Central PMCID: PMCPMC8646492.

219. Zhang X, Zou R, Liao X, Bernardo ABI, Du H, Wang Z, et al. Perceived Stress, Hope, and Health Outcomes Among Medical Staff in China During the COVID-19 Pandemic. Frontiers in Psychiatry. 2020;11. doi: 10.3389/fpsyt.2020.588008.

220. Zhang Y, Pi DD, Liu CJ, Li J, Xu F. Psychological impact of the COVID-19 epidemic among healthcare workers in paediatric intensive care units in China. PloS one. 2022;17(5 May). doi: 10.1371/journal.pone.0265377.

221. Zheng R, Zhou Y, Qiu M, Yan Y, Yue J, Yu L, et al. Prevalence and associated factors of depression, anxiety, and stress among Hubei pediatric nurses during COVID-19 pandemic. Comprehensive psychiatry. 2021;104. doi: 10.1016/j.comppsych.2020.152217.

222. Zhu Z, Xu S, Wang H, Liu Z, Wu J, Li G, et al. COVID-19 in Wuhan: Sociodemographic characteristics and hospital support measures associated with the immediate psychological impact on healthcare workers. EClinicalMedicine. 2020;24. doi: 10.1016/j.eclinm.2020.100443.

