## Supplementary material for "Mental health status and related factors influencing healthcare workers during the COVID-19 pandemic: a systematic review and meta-analysis": S1

**S1 File Retrieval strategy**

((("COVID-19"[Mesh]) OR ((((((((((((((((((((((((((((((((((((COVID 19[Title/Abstract]) OR (COVID 19[Title/Abstract])) OR (Infection, SARS-CoV-2[Title/Abstract])) OR (SARS CoV 2 Infection[Title/Abstract])) OR (SARS-CoV-2 Infections[Title/Abstract])) OR (2019 Novel Coronavirus Disease[Title/Abstract])) OR (2019 Novel Coronavirus Infection[Title/Abstract])) OR (2019-nCoV Disease[Title/Abstract])) OR (2019 nCoV Disease[Title/Abstract])) OR (2019-nCoV Diseases[Title/Abstract])) OR (Disease, 2019-nCoV[Title/Abstract])) OR (COVID-19 Virus Infection[Title/Abstract])) OR (COVID-19 Virus Infection[Title/Abstract])) OR (COVID-19 Virus Infections[Title/Abstract])) OR (Infection, COVID-19 Virus[Title/Abstract])) OR (Virus Infection, COVID-19[Title/Abstract])) OR (Coronavirus Disease 2019[Title/Abstract])) OR (Disease 2019, Coronavirus[Title/Abstract])) OR (Coronavirus Disease-19[Title/Abstract])) OR (Coronavirus Disease 19[Title/Abstract])) OR (Severe Acute Respiratory Syndrome Coronavirus 2 Infection[Title/Abstract])) OR (SARS Coronavirus 2 Infection[Title/Abstract])) OR (COVID-19 Virus Disease[Title/Abstract])) OR (COVID 19 Virus Disease[Title/Abstract])) OR (COVID-19 Virus Diseases[Title/Abstract])) OR (Disease, COVID-19 Virus[Title/Abstract])) OR (Virus Disease, COVID-19[Title/Abstract])) OR (2019-nCoV Infection[Title/Abstract])) OR (2019 nCoV Infection[Title/Abstract])) OR (2019-nCoV Infections[Title/Abstract])) OR (Infection, 2019-nCoV[Title/Abstract])) OR (COVID19[Title/Abstract])) OR (COVID-19 Pandemic[Title/Abstract])) OR (COVID 19 Pandemic[Title/Abstract])) OR (Pandemic, COVID-19[Title/Abstract])) OR (COVID-19 Pandemics[Title/Abstract])))

AND (("Mental Health"[Mesh]) OR (((Health, Mental[Title/Abstract]) OR (Mental Hygiene[Title/Abstract])) OR (Hygiene, Mental[Title/Abstract]))))

AND (((((((((((((Personnel, Health[Title/Abstract]) OR (Health Care Providers[Title/Abstract])) OR (Health Care Provider[Title/Abstract])) OR (Provider, Health Care[Title/Abstract])) OR (Healthcare Providers[Title/Abstract])) OR (Healthcare Provider[Title/Abstract])) OR (Provider, Healthcare[Title/Abstract])) OR (Healthcare Workers[Title/Abstract])) OR (Healthcare Worker[Title/Abstract])) OR (Health Care Professionals[Title/Abstract])) OR (Health Care Professional[Title/Abstract])) OR (Professional, Health Care[Title/Abstract])) OR ("Health Personnel"[Mesh]))
