## Supplementary material for "Mental health status and related factors influencing healthcare workers during the COVID-19 pandemic: a systematic review and meta-analysis": S3

**S3 File Reasons for exclusion during full- text screening**

**S2 Table: Reasons for exclusion during full- text screening**

| Author/year | Reason | Author/year | Reason |
| --- | --- | --- | --- |
| (Alonso et al. 2022) | Not outcome of interest | (Zhu et al., 2020) | Not population of interest |
| (Amerio et al. 2020) | Not outcome of interest | (Rana et al., 2020) | Not outcome of interest |
| (Antonijevic et al. 2020) | Not outcome of interest | (Restrepo-Martínez et al., 2021) | Not outcome of interest |
| (Firew et al., 2020) | Not outcome of interest | (Ristevska-Dimitrovska and Batic 2020) | Not population of interest |
| (Gupta et al., 2020) | letter to the editor | (Rossi et al., 2021) | Not outcome of interest |
| (Hendrickson et al., 2022) | Not outcome of interest | (Sangra et al., 2022) | Not outcome of interest |
| (Ilie et al. 2022) | Not outcome of interest | (Sarwar and Sarwar 2020) | letter to the editor |
| (Ishikawa et al., 2021) | Not outcome of interest | (Senczyszyn et al., 2020) | letter to the editor |
| (Lange et al., 2022) | Not outcome of interest | (Sil et al., 2020) | letter to the editor |
| (Liu et al., 2020b) | Not outcome of interest | (Smallwood and Willis 2021) | Not original article |
| (Lu et al., 2020) | Not outcome of interest | (Sonis et al., 2021) | letter to the editor |
| (Luceno-Moreno et al., 2022) | Not outcome of interest | (Spiller et al., 2022) | Not outcome of interest |
| (Mamani-Benito et al. 2021) | Not outcome of interest | (Styra et al., 2021) | Not population of interest |
| (Martin-Rodriguez et al., 2022) | Not outcome of interest | (Sumner 2022) | Not outcome of interest |
| (Mascayano et al., 2022) | Not outcome of interest | (Teng et al., 2020) | Not population of interest |
| (Mboua et al., 2021) | Not outcome of interest | (Topal et al., 2021) | Not outcome of interest |
| (Mo et al., 2021) | Not outcome of interest | (Usul et al., 2021) | Not outcome of interest |
| (Mohsin et al., 2021) | Not outcome of interest | (Wadoo et al., 2021) | letter to the editor |
| (Montoya et al. 2021) | Not outcome of interest | (Walvik et al., 2021) | Not population of interest |
| (Mustikasari et al., 2022) | Not outcome of interest | (Wozniak et al. 2021) | Not outcome of interest |
| (Mutair et al., 2021) | Not outcome of interest | (Wright et al., 2021) | Not population of interest |
| (Nadeem et al., 2021) | Not outcome of interest | (Wu and Wei, 2020) | Not outcome of interest |
| (Nicolaou et al. 2021) | Not outcome of interest | (Xu et al., 2022) | Not outcome of interest |
| (Northwood et al., 2021) | Not population of interest | (Yao et al. 2021) | Not outcome of interest |
| (Odikpo et al., 2021) | Not outcome of interest | (Yildirim et al. 2020) | Not outcome of interest |
| (Olivares-Tirado and Zanga-Pizarro, 2022) | Not outcome of interest | (Zandifar et al. 2020) | letter to the editor |
| (Orellano and Macavilca, 2020) | letter to the editor | (Zhang et al. 2022) | Not outcome of interest |
| (Ornell et al., 2020) | Not outcome of interest | (Zhang et al. 2021) | Not outcome of interest |
| (Zhu et al., 2020) | Not population of interest | (Zhou et al. 2021) | Not outcome of interest |
| (Quintana-Domeque et al., 2021) | Not outcome of interest |  |  |
