## Supplementary material for "Mental health status and related factors influencing healthcare workers during the COVID-19 pandemic: a systematic review and meta-analysis": S4

**S4 File Main characteristics of the included studies**

**S3 Table: Main characteristics of the included studies**

| Author/year | Country | Data time | Study design | Sample size | Male | Age | Healthcare workers | | | Burnout | | Anxity | | Depression | | ASD | | PTSD | |
| --- | --- | --- | --- | --- | --- | --- | --- | --- | --- | --- | --- | --- | --- | --- | --- | --- | --- | --- | --- |
|  |  |  |  |  |  |  | Doctor | Nurse | others | N(%) | scale | N(%) | scale | N(%) | scale | N(%) | scale | N(%) | scale |
| (Abu-Elenin, 2021) | Egypt | 2020.4-5 | cross-sectional study | 237 | 138 | 38.2 ± 6.2 | 237 |  |  |  |  | 79 | GAD-7 | 44 | PHQ-9 |  |  |  |  |
| (AlAteeq et al. 2020) | Saudi Arabia | 2020.3 | cross-sectional study | 502 | 342 | ≥18 | 111 | 132 | 259 |  |  | 51.4 | GAD-7 | 55.2 | PHQ-9 |  |  |  |  |
| (Ali et al. 2020) | Ireland | 2020.6 | array research | 472 | 146 | 40.7 | 91 | 137 | 244 |  |  | 45.1 | DASS-21 | 42.6 | DASS-21 |  |  | 41.3 | IES-R |
| (AlKandari et al. 2022) | Kuwait | 2020.7 | cross-sectional study | 378 | 117 | ≥18 |  |  |  |  |  | 52.9 | GAD-7 | 40.5 | PHQ-9 |  |  |  |  |
| (Alkhamees et al. 2021) | Saudi Arabia | 2020.3-4 | cross-sectional study | 121 | 70 | ≥24 |  |  | 121 | 27.3 | MBI |  |  | 27.3 | PHQ-9 |  |  |  |  |
| (Almalki et al. 2021) | Saudi Arabia | 2021.1-3 | cross-sectional study | 501 | 196 | ≥18 |  |  |  |  |  | 60.88 | DASS-21 | 54.69 | DASS-21 |  |  |  |  |
| (Alzahrani et al. 2022) | Saudi Arabia | 2021.9-12 | cross-sectional study | 251 | 74 | 21-54 |  | 251 |  |  |  | 43.4 | HADS | 34.7 | HADS |  |  |  |  |
| (Andlib et al. 2022) | Pakistan |  | cross-sectional study | 288 | 67 | 27.7 ± 4.4 |  | 288 |  | 48.6 | MBI |  |  |  |  |  |  |  |  |
| (Arshad and Islam 2020) | Pakistan | 2020.3 | cross-sectional study | 431 | 238 | ≥18 | 431 |  |  |  |  | 61.5 | GAD-7 |  |  |  |  |  |  |
| (Ashoor et al. 2021) | Saudi Arabia | 2020.6 | cross-sectional study | 129 | 82 | ≥25 | 129 |  |  |  |  |  |  | 75.2 | PHQ-9 |  |  |  |  |
| (Awano et al. 2020) | Japan | 2020.4-5 | cross-sectional study | 848 | 213 | 27-50 | 104 | 461 | 283 |  |  |  |  | 27.9 | GAD-7 |  |  |  |  |
| (Azizi et al. 2021) | Iran | 2020.4-5 | cross-sectional study | 7626 | 2044 | ≥20 | 1098 | 4409 | 2119 |  |  | 43 | DASS-21 | 45.8 | DASS-21 |  |  |  |  |
| (Azoulay et al. 2021) | France | 2020.10-12 | cross-sectional study | 845 | 274 | 28-41 | 175 | 412 | 258 |  |  | 60 | HADS | 36.1 | HADS |  |  | 28.4 | IES-R |
| (Babamiri et al. 2022) | Iran | 2020.5 | cross-sectional study | 942 | 390 | 20-60 | 76 | 86 | 780 | 50.2 | MBI | 62.2 | GHQ-28 | 17.5 | GHQ-28 |  |  |  |  |
| (Baminiwatta et al. 2021) | Sri Lanka | 2020.6-8 | cross-sectional study | 467 | 126 | ≥18 | 157 | 174 | 136 |  |  | 20.6 | DASS-21 | 19.5 | DASS-21 |  |  |  |  |
| (Cahill et al. 2022) | America | 2020.8-10 | cross-sectional study | 970 | 168 | 41.5± 11.1 |  | 379 |  |  |  | 80.6 | GAD-7 | 84.3 | PHQ-8 |  |  |  |  |
| (Çalişkan and Dost 2020) | Turkey | 2020.3 | cross-sectional study | 290 | 179 | 31.8±6.9 | 290 |  |  |  |  | 62 | HADS | 35.5 | HADS |  |  |  |  |
| (Carmassi et al. 2022) | Italy | 2020.5-6 | cross-sectional study | 139 | 60 | ≥18 |  |  | 139 |  |  | 31.7 | GAD-7 | 31.7 | PHQ-9 |  |  | 23 | IES-R |
| (Carriero et al. 2021) | Italy | 2020.4-6 | cross-sectional study | 770 | 198 | ≥18 | 95 | 675 |  |  |  | 59.4 | STAI | 11.8 | BDI |  |  |  |  |
| (Chatterjee et al. 2020) | India | 2020.3-4 | cross-sectional study | 152 | 119 | 42.05±12.19 | 152 |  |  |  |  | 39.5 | DASS-21 | 34.9 | DASS-21 |  |  |  |  |
| (Chatzittofis et al. 2021) | Cyprus | 2020.5 | cross-sectional study | 424 | 176 | 38.8±11.4 | 178 | 103 | 143 |  |  |  |  | 18.6 | PHQ-9 |  |  | 14.6 | IES-R |
| (Chen et al. 2021) | China | 2020.2 | cross-sectional study | 902 | 283 | 36.41 ± 8.56 | 543 | 311 | 48 |  |  | 47 | GAD-7 | 48.7 | PHQ-9 |  |  |  |  |
| (Chen, Lin, and Feng 2021) | China |  | cross-sectional study | 597 | 72 | ≥35 | 41 | 549 | 7 |  |  |  |  |  |  |  |  | 45.23 | IES-R |
| (Chen et al. 2021) | China | 2020.7-8 | cross-sectional study | 1083 | 41 | ≥18 |  | 1083 |  |  |  | 29.8 | DASS-21 | 22 | DASS-21 |  |  |  |  |
| (Chinvararak et al. 2022) | Thailand | 2021.5-6 | cross-sectional study | 986 | 107 | 34.89±11.05 | 50 | 623 | 313 |  |  | 33.1 | GAD-7 | 13.8 | PHQ-9 |  |  | 2.3 | DSM-5 |
| (Civantos et al. 2020) | Brazil | 2020.5 | cross-sectional study | 163 | 121 | ≥25 | 163 |  |  | 14.7 | IES-R | 16 | GAD-7 | 45.5 | PHQ-2 |  |  |  |  |
| (Çölkesen and Çölkesen 2021) | Turkey | 2020.5 | cross-sectional study | 435 | 208 | 34.34±8.34 | 240 | 160 | 33 |  |  | 43.4 | HADS | 65.1 | HADS |  |  |  |  |
| (Czepiel et al., 2022) | Netherlands | 2021.2-5 | cross-sectional study | 994 | 182 | 33-55 | 153 | 344 | 497 |  |  |  |  | 13 | PHQ-9 |  |  | 20 | DSM-5 |
| (d’Ussel et al., 2022) | France | 2020.7-10 | cross-sectional study | 780 | 142 |  | 125 | 367 | 288 |  |  | 41 | HADS | 21 | HADS |  |  | 14 | PCL |
| (Da'seh et al., 2022) | Jordan | 2021.8-10 | cross-sectional study | 364 | 137 | 30±3.53 |  | 364 |  |  |  | 48.9 | DASS-42 | 34.1 | DASS-42 |  |  | 67.3 | IES-R |
| (Dobson et al., 2021) | Australia | 2020.4-5 | cross-sectional study | 320 | 58 | ≥19 | 99 | 84 | 137 |  |  | 20 | GAD-7 | 21 | PHQ-9 |  |  | 29 | IES-R |
| (Etesam et al., 2022) | Iran | 2020.2-4 | cross-sectional study | 258 | 46 | 35.48±8.39 | 58 | 100 | 100 |  |  | 39.9 | GAD-2 | 39.1 | PHQ-2 |  |  |  |  |
| (Frajerman et al., 2022 | France | 2020.10-11 | cross-sectional study | 1529 | 625 | ≥20 | 1529 |  |  |  |  | 76.4 | HADS | 29 | HADS |  |  |  |  |
| (Fu et al., 2021) | China | 2020.2-3 | cross-sectional study | 7413 | 1244 | ≥18 | 1756 | 4368 | 1289 |  |  | 33.74 | GAD-7 | 27.65 | PHQ-9 |  |  |  |  |
| (GebreEyesus et al., 2021) | Ethiopia | 2020.11 | cross-sectional study | 322 | 167 | 28.71±5.88 | 44 | 113 | 165 |  |  | 36 | GAD-7 | 25.8 | PHQ-9 |  |  |  |  |
| (Gilleen et al., 2021) | United Kingdom | 2020.4-5 | cross-sectional study | 2773 | 395 |  | 386 | 852 | 1535 |  |  | 33.1 | GAD-7 | 28.1 | PHQ-9 |  |  | 60.6 | IES-R |
| (Goh et al., 2021) |  | 2020.3-6 | cross-sectional study | 3537 | 923 |  | 684 | 1590 | 1263 | 67 | OLBI | 20 | HADS | 11 | HADS |  |  |  |  |
| (Gonzalo et al., 2021) | Spain | 2020.5 | cross-sectional study | 1407 | 377 | 44.7±10.9 | 430 | 306 | 671 |  |  |  |  |  |  | 24.7 | SARS-Q |  |  |
| (Greenberg et al., 2021) | United Kingdom | 2020.6-7 | cross-sectional study | 709 |  |  | 291 | 344 | 74 |  |  | 11 | GAD-7 | 6 | PHQ-9 |  |  | 40 | PCL |
| (Guo et al., 2021) | China | 2020.5 | cross-sectional study | 1091 | 356 |  | 202 | 554 | 335 |  |  | 53 | GAD-7 | 56 | PHQ-9 |  |  | 11 | PCL |
| (Habtamu et al., 2021) | Ethiopia | 2020.5-6 | cross-sectional study | 238 | 101 | ≥18 | 59 | 164 | 15 |  |  | 31.1 | GAD-7 | 27.3 | PHQ-9 |  |  | 16 | PCL |
| (Hayat et al., 2021) | Pakistan | 2020.5-6 | cross-sectional study | 1094 | 371 | ≥20 | 742 | 277 | 75 |  |  | 33.3 | GAD-7 | 57.4 | PHQ-9 |  |  |  |  |
| (He et al., 2021) | China | 2020.1-5 | cross-sectional study | 1521 | 825 | 42.79±10.21 | 1521 |  |  |  |  | 11.11 | SAS | 16.9 | SDS |  |  |  |  |
| (Heesakkers et al., 2022) | Netherlands | 2020.9 | cross-sectional study | 589 | 159 | 44.8±11.9 |  | 589 |  |  |  | 29.9 | HADS | 21.1 | HADS |  |  | 17.7 | IES-R |
| (Hickling and Barnett, 2022) | America | 2020.7-9 | cross-sectional study | 112 | 9 |  |  | 112 |  |  |  | 62 | GAD-7 | 31 | PHQ-9 |  |  | 15 | PCL |
| (Hu et al., 2020) | China | 2020.1-2 | cross-sectional study | 2014 | 260 | 30.99±6.17 |  | 2014 |  | 60.5 | MBI | 25 | SAS | 91.2 | SDS |  |  |  |  |
| (Jakhar et al., 2021) | India | 2020.4 | cross-sectional study | 450 | 216 | 31.3±5.48 | 243 | 207 |  |  |  | 38.9 | DASS-21 | 43.6 | DASS-21 |  |  |  |  |
| (Jo et al., 2020) | South Korea |  | cross-sectional study | 253 | 43 | 39.1±12.3 | 27 | 149 | 77 |  |  |  |  |  |  |  |  | 21 | IES-R |
| (Kafle et al., 2021) | Nepal | 2020.7-9 | cross-sectional study | 270 | 122 | ≥20 | 59 | 57 | 154 |  |  | 41.4 | HADS | 24.1 | HADS |  |  |  |  |
| (Kamali et al., 2022) | Iran | 2020.4-5 | cross-sectional study | 7626 | 2044 | ≥21 | 1098 | 4409 | 2119 | 18.3 | MBI |  |  |  |  |  |  |  |  |
| (Kandouci et al., 2021) | Algeria | 2021.1-3 | cross-sectional study | 1005 | 245 | 27-38 | 518 |  | 487 |  |  | 23.8 | GAD-7 | 44.6 | CES-D |  |  |  |  |
| (Kapetanos et al., 2021) | Cyprus | 2020.5-6 | cross-sectional study | 381 | 76 |  | 49 | 277 | 55 | 12.3 | MBI | 28.6 | DASS-21 | 15 | DASS-21 |  |  |  |  |
| (Khan et al., 2021) | Canada | 2020.8-10 | cross-sectional study | 249 | 117 | 25-65 | 249 |  |  | 68 | MBI |  |  |  |  |  |  |  |  |
| (Khatun et al., 2021) | Bangladesh | 2020.5 | cross-sectional study | 114 | 76 |  | 114 |  |  |  |  | 32.5 | GAD-7 | 34.2 | PHQ-9 |  |  |  |  |
| (Kiliç et al., 2021) |  | 2020.4 | cross-sectional study | 544 | 200 | 36.85±10.46 | 275 | 231 | 38 |  |  | 60.1 | GAD-7 |  |  |  |  |  |  |
| (Lai et al. 2020) | China | 2020.1-2 | cross-sectional study | 1257 | 293 | ≥18 | 493 | 764 |  |  |  | 44.6 | GAD-7 | 50.4 | PHQ-9 |  |  |  |  |
| (Lamiani et al., 2021) | Italy | 2020.7-10 | cross-sectional study | 308 | 62 | 45.06±11.34 | 48 | 111 | 149 |  |  | 23 | STAI | 53 | HAM-D |  |  | 40 | PCL |
| (Lasalvia et al., 2021) | Italy | 2021.4-5 | cross-sectional study | 1033 | 234 |  | 309 | 379 | 345 | 40.6 | MBI | 55.7 | SAS | 40.6 | PHQ-9 |  |  |  |  |
| (Li et al., 2020a) | China | 2020.2 | cross-sectional study | 4369 | 0 | ≥19 | 159 | 1127 | 96 |  |  | 25.2 | GAD-7 | 14.2 | PHQ-9 | 31.6 | IES-R |  |  |
| (Li et al., 2022) | China | 2022.1 | cross-sectional study | 256 | 77 | ≥21 | 256 |  |  |  |  | 37.1 | DASS-42 | 33.2 | DASS-42 |  |  |  |  |
| (Li et al., 2021) | China | 2020.2-3 | cross-sectional study | 318 | 3 | 35.13±8.65 |  | 318 |  |  |  | 30.19 | SCL-90 | 29.25 | SCL-90 |  |  |  |  |
| (Li et al., 2020b) | China |  | cross-sectional study | 5331 |  |  |  |  | 5331 |  |  | 6.1 | GAD-7 | 6.5 | PHQ-9 |  |  |  |  |
| (Li et al., 2022c) | China | 2021.6-7 | cross-sectional study | 138279 | 5885 |  |  | 138279 |  |  |  | 41.8 | SAS | 55.5 | SDS |  |  | 34 | MBI-HAA |
| (Liu et al., 2020a) | China | 2020.2 | cross-sectional study | 512 | 79 | ≥18 |  |  |  |  |  | 12.5 | SAS |  |  |  |  |  |  |
| (Lixia et al., 2022) | China | 2020.6 | cross-sectional study | 33706 | 7846 | ≥18 | 13803 | 15324 | 4579 |  |  | 24.4 | GAD-7 | 35.8 | PHQ-9 |  |  | 5 | PCL |
| (Luceño-Moreno et al., 2020) | Spain | 2020.4 | cross-sectional study | 1422 | 194 | 43.88±10.82 |  |  |  |  |  | 58.6 | HADS | 46 | HADS |  |  | 56.6 | IES-R |
| (Martin et al., 2021) | Spain | 2020.4-9 | array research | 2089 | 398 |  |  |  |  |  |  | 51.75 | GAD-7 | 38.58 | PHQ-9 |  |  |  |  |
| (Martínez Pajuelo et al., 2022) | Peru | 2021.4-7 | cross-sectional study | 109 | 47 |  | 44 | 35 | 29 |  |  | 32.11 | GAD-7 | 30.28 | PHQ-9 |  |  | 31.19 | DSM-5 |
| (Martínez-Caballero et al., 2021) | Spain | 2020.5-7 | cross-sectional study | 317 | 43 |  | 61 | 78 | 178 |  |  |  |  |  |  |  |  | 30.9 | DTS-8 |
| (Martsenkovskyi et al. 2022) | Ukraine | 2021.4-5 | cross-sectional study | 392 | 65 | ≥20 | 281 |  |  |  |  | 55.4 | DASS-21 | 50.5 | DASS-21 |  |  | 20 | DSM-5 |
| (Mattila et al., 2021) | Finland | 2020.4-5 | cross-sectional study | 1995 | 255 | ≥18 |  |  |  |  |  | 45 | DASS-21 |  |  |  |  |  |  |
| (McGuinness et al., 2022) | Australia | 2021.5-7 | array research | 984 | 267 |  |  |  |  |  |  | 14 | DASS-21 | 22.5 | PHQ-9 |  |  | 20.4 | IES-R |
| (Mekhemar et al., 2021a) | Germany | 2020.7-2021.1 | cross-sectional study | 252 | 5 | ≥18 |  | 252 |  |  |  | 36.1 | DASS-42 | 43.7 | DASS-21 |  |  |  |  |
| (Mekhemar et al., 2021b) | Germany | 2020.7-11 | cross-sectional study | 732 | 293 | ≥18 | 732 |  |  |  |  | 43.3 | DASS-21 | 30.6 | DASS-21 |  |  |  |  |
| (Mennicken et al. 2022) | Belgium | 2020.6-7 | cross-sectional study | 542 | 108 | 23-49 | 146 | 396 |  |  |  | 55 | HADS | 32 | HADS |  |  | 47 | IES-R |
| (Meo et al., 2021) | Saudi Arabia | 2020.7-12 | cross-sectional study | 1678 | 819 | ≥20 | 447 | 259 | 972 |  |  | 58.9 | GAD-7 |  |  |  |  |  |  |
| (Mi et al. 2021) | China | 2020.4 | cross-sectional study | 1029 | 395 | 19-78 | 268 | 219 | 542 |  |  | 6.61 | PHQ-4 | 13.31 | PHQ-4 |  |  |  |  |
| (Morawa et al., 2021) | Germany | 2020.4-7 | cross-sectional study | 3678 | 927 |  | 1061 | 1275 | 1342 |  |  | 19.1 | GAD-2 | 20.9 | PHQ-2 |  |  |  |  |
| (Moro et al., 2022) | Italy | 2020.5-7 | cross-sectional study | 4320 | 1297 |  | 1535 | 542 | 2243 |  |  |  |  | 7.55 | PHQ-9 |  |  |  |  |
| (Mosheva et al., 2021) | Israel | 2020.4 | cross-sectional study | 828 | 271 | 41.7±11.1 | 349 | 479 |  |  |  | 32.9 | PROMIS | 19.3 | PHQ-9 |  |  | 13.9 | DSM-5 |
| (Motahedi et al., 2021) | Iran | 2020.5-8 | cross-sectional study | 140 | 45 | 34.24±7.31 |  |  |  |  |  | 22.9 | GAD-7 | 57.6 | CES-D |  |  |  |  |
| (Mulatu et al., 2021) | Ethiopia | 2020.8 | cross-sectional study | 420 | 246 | ≥18 | 115 | 237 | 68 |  |  | 21.9 | GAD-7 | 20.2 | PHQ-9 |  |  |  |  |
| (Nagarkar et al., 2022) | India | 2020.5 | cross-sectional study | 344 | 190 |  |  |  |  |  |  | 18 | GAD-7 | 22 | PHQ-9 |  |  |  |  |
| (Nguyen et al., 2021) | Vietnam | 2020.4 | cross-sectional study | 349 | 136 | 35.2±8.8 | 199 | 82 | 68 |  |  |  |  |  |  |  |  | 4.6 | IES-R |
| (Ning et al., 2020) | China | 2020.2 | cross-sectional study | 612 | 166 |  | 317 | 295 |  |  |  | 16.3 | SAS | 25 | SDS |  |  |  |  |
| (Nordin et al., 2022) | Malaysia | 2020.7-8 | cross-sectional study | 981 | 250 |  |  |  |  |  |  | 17.1 | DASS-21 | 8.4 | DASS-21 |  |  |  |  |
| (NS et al., 2021) | Saudi Arabia | 2020.4-6 | cross-sectional study | 554 | 165 |  | 214 | 298 | 42 |  |  |  |  | 48 | PHQ-9 |  |  |  |  |
| (Osório et al., 2021) | Brazil | 2020.5-8 | cross-sectional study | 916 | 186 | 35.2±9.2 | 275 | 376 | 265 |  |  | 43.3 | GAD-7 | 40.2 | PHQ-9 |  |  | 36 | PCL |
| (Oteir et al., 2022) | Jordan | 2020.5-6 | cross-sectional study | 122 | 98 | 32.1±5.8 | 54 | 40 | 28 |  |  | 33.6 | GAD-7 | 40.2 | PHQ-9 |  |  |  |  |
| (Pan et al., 2022) | China | 2020.2-12 | cross-sectional study | 194 | 36 |  | 42 | 148 | 4 |  |  | 32.5 | GAD-7 | 37.6 | PHQ-9 |  |  |  |  |
| (Pandey et al. 2021) | Nepal | 2020.4-5 | cross-sectional study | 404 | 257 | 32.25 ± 8.23 | 154 | 188 | 62 |  |  | 35.6 | DASS-21 | 28.9 | DASS-21 |  |  |  |  |
| (Paniagua-Ávila et al., 2022 | Guatemala | 2020.7-9 | cross-sectional study | 1522 | 508 | ≥18 | 566 | 326 | 630 |  |  |  |  | 23 | PHQ-9 |  |  |  |  |
| (Pappa et al., 2021) | Greece | 2020.5-6 | cross-sectional study | 464 | 145 | 41.37±11 | 179 | 200 | 85 | 65 | MBI | 25 | GAD-7 | 30 | PHQ-9 |  |  | 33 | IES-R |
| (Parthasarathy et al., 2021) | India | 2020.7-9 | cross-sectional study | 3083 | 1420 | 36±8.3 | 826 | 504 | 1753 |  |  | 26.6 | GAD-2 | 23.8 | PHQ-4 |  |  |  |  |
| (Pelissier et al., 2021) | France | 2020.5-6 | cross-sectional study | 340 | 80 |  |  |  | 340 |  |  | 32.1 | HADS | 7.65 | HADS |  |  |  |  |
| (Pérez Herrera et al. 2021) | Colombia | 2020.10-11 | cross-sectional study | 133 |  |  | 133 |  |  |  |  | 63.91 | GAD-7 | 33.83 | PHQ-9 |  |  |  |  |
| (Pisanu et al., 2022) | Italy | 2020.4-5 | cross-sectional study | 576 | 184 | 22-69 | 223 | 353 |  |  |  | 76.5 | PROMIS |  |  |  |  |  |  |
| (Proserpio et al., 2022) | Italy | 2020.6-7 | cross-sectional study | 964 | 197 | ≥20 |  |  |  |  |  | 69.7 | STAI | 32.8 | BDI-II |  |  |  |  |
| (Rahman et al., 2021) | Bangladesh | 2020.8-10 | cross-sectional study | 347 | 221 |  | 347 |  |  |  |  | 35.2 | DASS-21 | 55.3 | DASS-21 |  |  |  |  |
| (Repon et al., 2021) | Bangladesh | 2020.7-9 | cross-sectional study | 355 | 204 | 20-60 | 106 | 91 | 158 |  |  | 78 | GAD-7 | 44 | PHQ-9 |  |  |  |  |
| (Riaz et al., 2021) | Pakistan | 2020.7 | cross-sectional study | 134 | 85 | 21-70 | 66 | 24 | 44 |  |  | 6 | DASS-21 | 15.7 | DASS-21 |  |  |  |  |
| (RilleraMarzo et al., 2021) | Iran | 2020.4-5 | cross-sectional study | 516 | 212 | 25-60 | 105 | 8 | 403 |  |  | 70.74 | GAD-7 | 50.97 | PHQ-9 |  |  |  |  |
| (Rossi et al. 2020) | Italy | 2020.3 | cross-sectional study | 1379 | 315 | 39 ± 16 | 433 | 472 | 474 |  |  | 19.8 | GAD-7 | 24.73 | PHQ-9 |  |  |  |  |
| (Sabbaghi et al., 2022) | Iran | 2021.8-9 | cross-sectional study | 544 | 512 | 34.1±7.4 |  |  | 544 |  |  | 36.7 | DASS-21 | 36.2 | DASS-21 |  |  |  |  |
| (Salehiniya and Abbaszadeh, 2021) | Iran | 2020.5 | cross-sectional study | 320 | 172 | 44.38±10.66 | 320 |  |  |  |  | 32.5 | GHQ-28 |  |  |  |  |  |  |
| (Salman et al., 2022) | Pakistan | 2020.4-5 | cross-sectional study | 398 | 183 | 28.67±4.15 | 205 | 133 | 60 |  |  | 21.4 | GAD-7 | 21.9 | PHQ-9 |  |  |  |  |
| (Samir AlKudsi et al., 2022) | Qatar | 2020.10-2021.2 | cross-sectional study | 256 | 144 | 35.8±7.5 |  |  | 256 | 44.4 | MBI | 53.2 | DASS-21 | 44.8 | DASS-21 |  |  |  |  |
| (Sertoz et al., 2021) | Turkey | 2020.4 | cross-sectional study | 683 | 208 | 20-66 | 94 | 200 | 389 |  |  | 70 | HADS | 38.5 | HADS |  |  |  |  |
| (Setiawati et al., 2021) | Indonesia | 2020.6 | cross-sectional study | 227 | 38 | 39.67±9.434 | 2 | 134 | 91 |  |  | 33 | STAI |  |  |  |  |  |  |
| (Shah et al., 2021a) | America | 2020.5-6 | cross-sectional study | 153 | 114 | 46.1±10.1 | 153 |  |  |  |  | 7.2 | GAD-7 | 8.5 | PHQ-8 |  |  |  |  |
| (Shah et al., 2021) | Kenya | 2020.8-11 | cross-sectional study | 433 | 176 |  | 243 | 190 |  | 45.8 | SPFI | 44.3 | GAD-7 | 53.6 | PHQ-9 |  |  |  |  |
| (Shah et al., 2020) | United Kingdom |  | cross-sectional study | 207 | 39 | 20-69 | 157 |  | 50 |  |  | 16 | PHQ-2 | 24.64 | GAD-2 |  |  |  |  |
| (Shechter et al., 2022) | America | 2020.4 | cross-sectional study | 230 | 46 | 31-48 | 50 | 115 | 65 |  |  |  |  |  |  |  |  | 59.1 | PC-PTSD |
| (Shechter et al., 2020) | America | 2020.4 | cross-sectional study | 657 | 143 | ≥18 | 141 | 313 | 203 |  |  | 33 | GAD-2 | 48 | PHQ-2 |  |  |  |  |
| (Shen et al., 2020) | China | 2020.1-2 | cross-sectional study | 373 | 48 | 29.55±6.93 | 98 | 275 |  |  |  | 5.63 | SCL-90 | 4.29 | SCL-90 |  |  |  |  |
| (Shrestha, 2020) | Nepal | 2020.4-5 | cross-sectional study | 101 | 43 |  | 60 | 41 |  |  |  | 73.3 | GAD-7 |  |  |  |  |  |  |
| (Siamisang et al., 2022) | Botswana | 2021.7-9 | cross-sectional study | 447 | 176 | 20-67 | 32 | 156 | 259 |  |  | 28.2 | DASS-42 | 21 | DASS-42 |  |  |  |  |
| (Simonetti et al., 2021) | Italy | 2020.2-4 | cross-sectional study | 1005 | 342 | 40.2±10.8 |  | 1005 |  |  |  | 33.23 | SAS |  |  |  |  |  |  |
| (Singh et al., 2021) | India | 2020.7-8 | cross-sectional study | 348 | 194 | 31.8±7.1 | 242 | 106 |  |  |  | 44.3 | GAD-7 | 54 | PHQ-9 |  |  |  |  |
| (Sitanggang et al., 2021) | Indonesia | 2020.7 | cross-sectional study | 635 | 288 | 32.7±7.7 |  |  | 635 |  |  | 19.7 | DASS-21 | 13.2 | DASS-21 |  |  |  |  |
| (Smallwood et al., 2021) | Australia | 2020.8-10 | cross-sectional study | 7846 | 1458 | ≥20 | 2436 | 3088 | 2322 | 70.9 | MBI | 59.8 | GAD-7 | 57.3 | PHQ-9 |  |  | 5.4 | IES-R |
| (Sobregrau Sangra et al., 2022) | Spain | 2020.7-10 | cross-sectional study | 184 | 28 |  | 43 | 104 | 37 |  |  | 90 | STAI | 13 | PHQ-2 |  |  | 23 | PCL |
| (Song et al., 2020) | China | 2020.2-3 | cross-sectional study | 14825 | 5289 | ≥18 | 6093 | 8732 |  |  |  |  |  | 25.2 | CES-D |  |  | 9.1 | DSM-5 |
| (Srikanth et al., 2022) | America | 2021.8-9 | cross-sectional study | 123 | 70 | ≥18 |  |  | 123 |  |  |  |  | 67.5 | PHQ-2 |  |  |  |  |
| (Stocchetti et al., 2021) | Italy | 2021.1 | cross-sectional study | 134 | 55 |  | 52 | 82 |  | 60 | MBI | 53 | HADS | 45 | HADS |  |  |  |  |
| (Su et al. 2021) | China | 2020.7-8 | cross-sectional study | 503 | 77 | ≥21 | 21 | 198 | 284 |  |  | 39.6 | DASS-21 | 31.2 | DASS-21 |  |  |  |  |
| (Su et al. 2021) | China | 2020.2 | cross-sectional study | 2920 | 385 | ≥18 |  |  |  |  |  | 27.1 | HADS | 22.2 | HADS |  |  |  |  |
| (Sun et al. 2020) | China | 2020.1-2 | cross-sectional study | 442 | 74 | ≥18 | 53 |  | 389 |  |  |  |  |  |  |  |  |  |  |
| (Suryavanshi et al. 2020) | India | 2020.5 | cross-sectional study | 197 | 96 | ≥18 | 66 | 47 | 44 |  |  | 22 | GAD-7 | 29 | PHQ-9 |  |  |  |  |
| (Syamlan et al. 2022) | Indonesia | 2020.12-2021.2 | cross-sectional study | 392 | 127 | 33.5±9.4 | 227 | 52 | 113 |  |  | 29.4 | DASS-21 | 44.9 | DASS-21 |  |  |  |  |
| (Tan et al., 2020) | Singapore | 2020.2-3 | cross-sectional study | 470 | 149 |  | 135 | 161 | 174 |  |  | 14.5 | DASS-21 | 8.9 | DASS-21 |  |  | 7.7 | IES-R |
| (Tan et al. 2021) |  | 2020.6 | cross-sectional study | 3391 | 2377 | ≥21 |  |  |  |  |  | 30.8 | DASS-21 | 32.8 | DASS-21 |  |  | 24 | IES-R |
| (Tang et al., 2022) | China | 2022.1-2 | cross-sectional study | 388 | 177 | ≥20 | 222 | 74 | 92 |  |  | 25.3 | GAD-7 | 40.7 | PHQ-9 |  |  |  |  |
| (Tao et al. 2021) | China | 2020.4 | cross-sectional study | 969 | 310 | 18-65 |  |  |  |  |  | 7.1 | GAD-7 | 13.8 | PHQ-9 |  |  | 8.5 | PSS-10 |
| (Taşdelen et al., 2022) | Turkey | 2020.4-5 | cross-sectional study | 634 | 202 | 35.89±8.63 | 426 | 118 | 90 |  |  | 35 | DASS-21 | 36 | DASS-21 |  |  |  |  |
| (Than et al., 2020) | Vietnam | 2020.3-4 | cross-sectional study | 173 | 55 | 27-36 | 43 | 109 | 21 |  |  | 33.5 | DASS-21 | 20.2 | DASS-21 |  |  |  |  |
| (Turan et al., 2022) | Turkey | 2020.4 | cross-sectional study | 300 | 193 | ≥20 | 246 | 33 | 21 |  |  | 44.6 | HADS | 68.2 | HADS |  |  |  |  |
| (Urzua et al., 2020) | Chile |  | cross-sectional study | 125 |  | 18-67 | 32 | 22 | 71 |  |  | 74 | GAD-7 | 65 | PHQ-9 |  |  |  |  |
| (Uyaroğlu et al., 2020) | Turkey | 2020.4 | cross-sectional study | 113 | 60 |  |  |  |  |  |  | 49.6 | GAD-7/BAI |  |  |  |  |  |  |
| (Uz et al., 2022) | Turkey | 2020.6-8 | cross-sectional study | 221 | 69 | 20-66 | 64 | 87 | 70 |  |  | 26.2 | HADS | 39.8 | HADS |  |  |  |  |
| (Vadi et al., 2022) | India | 2020.12-2021.1 | cross-sectional study | 153 | 64 | ≥21 |  |  |  |  |  | 16.3 | SAS | 28.1 | BDI |  |  | 21.6 | PSS-10 |
| (Valaine et al., 2021) | Latvia | 2020.4-6 | cross-sectional study | 844 | 127 |  | 350 | 384 | 110 |  |  | 17.2 | GAD-7 | 24.8 | PHQ-9 |  |  |  |  |
| (van de Venter et al., 2021) | South Africa | 2020.6-8 | cross-sectional study | 248 | 25 |  |  |  | 248 |  |  | 69.8 | CAS |  |  |  |  |  |  |
| (Van Wert et al., 2022) | America | 2020.9-11 | cross-sectional study | 605 | 108 | ≥19 |  |  |  |  |  | 14.2 | GAD-7 | 43.1 | PHQ-2 |  |  | 22.3 | IES-R |
| (Vancappel et al., 2021) | France | 2020.3-6 | cross-sectional study | 1010 | 172 | 39.24±11.13 | 185 | 360 | 465 | 66.6 | MBI |  |  |  |  |  |  | 57.8 | IES-R |
| (Wanigasooriya et al., 2020) | United Kingdom | 2020.6-7 | cross-sectional study | 2638 | 524 |  | 460 | 775 | 1403 |  |  | 34.3 | PHQ-4 | 31.2 | PHQ-4 |  |  | 24.5 | IES-R |
| (Wayessa et al., 2021) | Ethiopia | 2020.6-7 | cross-sectional study | 275 | 173 | 29.83±4.79 | 4 | 98 | 173 |  |  |  |  | 21.5 | DASS-21 |  |  |  |  |
| (Weibelzahl et al., 2021) | Germany | 2020.5-7 | cross-sectional study | 300 | 57 | 40.65±12.9 |  |  |  |  |  | 41 | ISR | 74 | IRS |  |  |  |  |
| (Xiong et al., 2020) | China | 2020.2 | cross-sectional study | 223 | 6 | ≤55 |  | 223 |  |  |  | 40.8 | GAD-7 | 26.4 | PHQ-9 |  |  |  |  |
| (Yadeta et al., 2021) | Ethiopia | 2020.10-11 | cross-sectional study | 265 | 149 | 29.29±6.4 | 18 | 188 | 59 |  |  |  |  | 66.4 | PHQ-9 |  |  |  |  |
| (Yang et al., 2022) | China | 2020.3-5 | cross-sectional study | 1993 | 337 | ≥20 | 396 | 1197 | 400 |  |  |  |  |  |  |  |  | 9.3 | DSM-5 |
| (Youssfi et al. 2021) | Tunisia | 2020.4-5 | cross-sectional study | 105 | 28 | ≥20 | 105 |  |  | 73.3 | ProQOL | 64.8 | GAD-7 |  |  |  |  |  |  |
| (Yu et al. 2022) | America | 2020.3 | cross-sectional study | 889 | 239 | ≥18 |  |  |  | 25 | ProQoL | 6 | CAS |  |  |  |  | 13 | DSM-5 |
| (Zarzour et al. 2022) | Lebanon | 2020.4 | cross-sectional study | 618 | 117 | ≥18 | 115 | 503 |  |  |  | 61.5 | STAI |  |  |  |  |  |  |
| (Zhan et al. 2022) | China | 2020.2-3 | cross-sectional study | 201 | 51 | 33.31±7.12 | 127 | 74 |  |  |  | 15.9 | SAS | 21.9 | SDS |  |  |  |  |
| (Zhang et al. 2020) | China | 2020.6 | cross-sectional study | 642 | 96 | ≥18 | 174 | 468 |  |  |  | 30.71 | HADS | 71.26 | HADS |  |  | 20.87 | PCL |
| (Zhang et al. 2021) | China | 2020.2 | cross-sectional study | 1040 | 215 | ≥18 |  |  | 1040 |  |  |  |  | 44.2 | SAS |  |  |  |  |
| (Zhang et al. 2020) | China | 2020.2-3 | cross-sectional study | 319 | 121 | 30.42±5.16 | 113 | 149 | 57 |  |  | 29.7 | HADS | 28.8 | HADS |  |  |  |  |
| (Zhang et al. 2022) | China | 2020.4 | cross-sectional study | 2109 | 316 | 32.42±6.66 | 739 | 1370 |  |  |  | 36.46 | DASS-21 | 39.69 | DASS-21 |  |  | 45.99 | IES-R |
| (Zheng et al. 2021) | China | 2020.3 | cross-sectional study | 617 | 3 | ≥18 |  | 617 |  |  |  | 32.6 | DASS-21 | 15.4 | DASS-21 |  |  |  |  |
| (Zhu et al. 2020) | China | 2020.2 | cross-sectional study | 5062 | 758 | 18-64 | 1004 | 3417 | 641 |  |  | 24.1 | GAD-7 | 13.5 | PHQ-9 |  |  |  |  |
