## Supplementary material for "Mental health status and related factors influencing healthcare workers during the COVID-19 pandemic: a systematic review and meta-analysis": S5

**S5 File Quality assessment**

**S4 Table: American Institute for Healthcare Quality and Research (AHRQ) cross-sectional study evaluation criteria**

| Author/year | Q1 | Q2 | Q3 | Q4 | Q5 | Q6 | Q7 | Q8 | Q9 | Q10 | Q11 | Score | Quality |
| --- | --- | --- | --- | --- | --- | --- | --- | --- | --- | --- | --- | --- | --- |
| (Abu-Elenin, 2021) | Y | N | Y | Y | Y | N | N | N | N | Y | N | 5 | M |
| (AlAteeq et al. 2020) | Y | N | Y | Y | Y | N | N | Y | N | N | N | 5 | M |
| (AlKandari et al. 2022) | Y | N | Y | Y | Y | N | N | Y | N | N | N | 5 | M |
| (Alkhamees et al. 2021) | Y | N | Y | Y | Y | N | N | Y | N | Y | N | 6 | M |
| (Almalki et al. 2021) | Y | N | Y | Y | Y | N | N | Y | N | N | N | 5 | M |
| (Alzahrani et al. 2022) | Y | N | Y | Y | Y | N | N | N | N | Y | N | 5 | M |
| (Andlib et al. 2022) | Y | Y | N | Y | Y | N | N | Y | N | N | N | 5 | M |
| (Arshad and Islam 2020) | Y | N | Y | Y | Y | N | Y | N | N | Y | N | 6 | M |
| (Ashoor et al. 2021) | Y | N | Y | Y | Y | N | Y | N | N | Y | N | 6 | M |
| (Awano et al. 2020) | Y | N | Y | Y | Y | N | N | N | N | Y | N | 5 | M |
| (Azizi et al. 2021) | Y | Y | Y | Y | Y | N | N | N | Y | N | N | 6 | M |
| (Azoulay et al. 2021) | Y | N | Y | Y | Y | N | N | Y | N | N | N | 5 | M |
| (Babamiri et al. 2022) | Y | N | Y | Y | Y | N | N | N | N | Y | N | 5 | M |
| (Baminiwatta et al. 2021) | Y | N | Y | Y | Y | N | Y | Y | N | N | N | 6 | M |
| (Cahill et al. 2022) | Y | N | Y | Y | Y | Y | N | Y | N | Y | N | 7 | M |
| (Çalişkan and Dost 2020) | Y | N | Y | Y | Y | N | N | N | N | Y | N | 5 | M |
| (Carmassi et al. 2022) | Y | N | Y | Y | Y | Y | N | N | N | N | N | 5 | M |
| (Carriero et al. 2021) | Y | N | Y | Y | Y | N | Y | N | N | N | N | 5 | M |
| (Chatterjee et al. 2020) | Y | N | Y | Y | Y | N | N | Y | N | N | N | 5 | M |
| (Chatzittofis et al. 2021) | Y | N | Y | Y | Y | N | N | Y | N | N | N | 5 | M |
| (Chen et al. 2021) | Y | N | Y | Y | Y | N | N | Y | N | N | N | 5 | M |
| (Chen, Lin, and Feng 2021) | Y | N | Y | Y | Y | N | Y | Y | N | N | N | 6 | M |
| (Chen et al. 2021) | Y | N | Y | Y | Y | N | N | Y | N | N | N | 5 | M |
| (Chinvararak et al. 2022) | Y | N | Y | Y | Y | N | N | N | N | Y | N | 5 | M |
| (Civantos et al. 2020) | Y | N | Y | Y | Y | N | N | N | N | N | N | 4 | M |
| (Çölkesen and Çölkesen 2021) | Y | N | Y | Y | Y | N | N | N | N | N | N | 4 | M |
| (Czepiel et al., 2022) | Y | Y | Y | Y | Y | N | Y | Y | N | N | N | 7 | M |
| (d’Ussel et al., 2022) | Y | N | Y | Y | Y | N | N | Y | N | Y | N | 6 | M |
| (Da'seh et al., 2022) | Y | Y | Y | Y | Y | N | N | Y | N | Y | N | 7 | M |
| (Dobson et al., 2021) | Y | N | Y | Y | Y | N | Y | Y | N | Y | N | 7 | M |
| (Etesam et al., 2022) | Y | Y | Y | Y | Y | N | N | Y | Y | Y | N | 8 | H |
| (Frajerman et al., 2022 | Y | N | Y | Y | Y | N | N | Y | N | N | N | 5 | M |
| (Fu et al., 2021) | Y | Y | Y | Y | Y | N | N | Y | N | Y | N | 7 | M |
| (GebreEyesus et al., 2021) | Y | Y | Y | Y | Y | Y | N | Y | N | Y | N | 8 | H |
| (Gilleen et al., 2021) | Y | N | Y | Y | Y | N | N | Y | N | Y | N | 6 | M |
| (Goh et al., 2021) | Y | N | Y | Y | Y | N | N | Y | N | N | N | 5 | M |
| (Gonzalo et al., 2021) | Y | Y | Y | Y | Y | N | N | Y | N | Y | N | 7 | M |
| (Greenberg et al., 2021) | Y | N | Y | Y | Y | N | N | Y | N | N | N | 5 | M |
| (Guo et al., 2021) | Y | Y | Y | Y | Y | N | N | Y | N | Y | N | 7 | M |
| (Habtamu et al., 2021) | Y | N | Y | Y | Y | N | N | Y | N | Y | N | 6 | M |
| (Hayat et al., 2021) | Y | N | Y | Y | Y | N | N | Y | N | N | N | 5 | M |
| (He et al., 2021) | Y | N | Y | Y | Y | N | N | Y | N | N | N | 5 | M |
| (Heesakkers et al., 2022) | Y | N | Y | Y | Y | N | N | Y | N | Y | N | 6 | M |
| (Hickling and Barnett, 2022) | Y | N | Y | Y | Y | N | N | Y | N | N | N | 5 | M |
| (Hu et al., 2020) | Y | Y | Y | Y | Y | Y | N | Y | N | Y | N | 8 | H |
| (Jakhar et al., 2021) | Y | N | Y | Y | Y | N | Y | Y | N | Y | N | 7 | M |
| (Jo et al., 2020) | Y | N | Y | Y | Y | N | N | Y | N | N | N | 5 | M |
| (Kafle et al., 2021) | Y | Y | Y | Y | Y | N | N | N | N | Y | N | 6 | M |
| (Kamali et al., 2022) | Y | N | Y | Y | Y | N | N | Y | Y | Y | N | 7 | M |
| (Kandouci et al., 2021) | Y | Y | Y | Y | Y | N | N | Y | N | Y | N | 7 | M |
| (Kapetanos et al., 2021) | Y | Y | Y | Y | Y | Y | N | Y | N | Y | N | 8 | H |
| (Khan et al., 2021) | Y | N | Y | Y | Y | N | Y | Y | Y | Y | N | 8 | H |
| (Khatun et al., 2021) | Y | Y | Y | Y | Y | N | N | Y | N | Y | N | 7 | M |
| (Kiliç et al., 2021) | Y | N | Y | Y | Y | N | N | N | N | N | N | 4 | M |
| (Lai et al. 2020) | Y | N | Y | Y | Y | N | N | Y | N | Y | N | 6 | M |
| (Lamiani et al., 2021) | Y | N | Y | Y | Y | N | N | Y | N | Y | N | 6 | M |
| (Lasalvia et al., 2021) | Y | N | Y | Y | Y | N | N | Y | N | Y | N | 6 | M |
| (Li et al., 2020) | Y | N | Y | Y | Y | N | N | Y | N | Y | N | 6 | M |
| (Li et al., 2022) | Y | Y | Y | Y | Y | N | N | Y | N | Y | N | 7 | M |
| (Li et al., 2021) | Y | N | Y | Y | Y | N | N | Y | N | Y | N | 6 | M |
| (Li et al., 2020b) | Y | N | Y | Y | Y | N | N | Y | N | Y | N | 6 | M |
| (Li et al., 2022c) | Y | N | Y | Y | Y | Y | N | Y | N | Y | N | 7 | M |
| (Liu et al., 2020) | Y | N | Y | Y | Y | N | N | Y | N | Y | N | 6 | M |
| (Lixia et al., 2022) | Y | Y | Y | Y | Y | N | N | N | N | N | N | 5 | M |
| (Luceño-Moreno et al., 2020) | Y | Y | Y | Y | Y | N | N | Y | N | Y | N | 7 | M |
| (Martínez Pajuelo et al., 2022) | Y | Y | Y | Y | Y | N | Y | Y | N | Y | N | 8 | H |
| (Martínez Pajuelo et al., 2022) | Y | Y | Y | Y | Y | N | N | N | N | Y | N | 6 | M |
| (Martsenkovskyi et al. 2022) | Y | N | Y | Y | Y | N | Y | Y | N | Y | N | 7 | M |
| (Mattila et al., 2021) | Y | N | Y | Y | Y | Y | N | Y | N | N | N | 6 | M |
| (Mekhemar et al., 2021a) | Y | N | Y | Y | Y | N | N | Y | N | Y | N | 6 | M |
| (Mekhemar et al., 2021b) | Y | N | Y | Y | Y | N | N | Y | N | Y | N | 6 | M |
| (Mennicken et al., 2022) | Y | N | Y | Y | Y | N | N | Y | N | Y | N | 6 | M |
| (Meo et al., 2021) | Y | Y | Y | Y | Y | N | Y | N | N | Y | N | 7 | M |
| (Mi et al. 2021) | Y | Y | Y | Y | Y | N | Y | Y | N | N | N | 7 | M |
| (Morawa et al., 2021) | Y | Y | Y | Y | Y | N | N | Y | N | Y | N | 7 | M |
| (Moro et al., 2022) | Y | Y | Y | Y | Y | N | Y | Y | N | Y | N | 8 | H |
| (Mosheva et al., 2021) | Y | N | Y | Y | Y | N | N | Y | N | N | N | 5 | M |
| (Motahedi et al., 2021) | Y | Y | Y | Y | Y | Y | N | Y | N | Y | N | 8 | H |
| (Mulatu et al., 2021) | Y | Y | Y | Y | Y | Y | N | Y | N | Y | N | 8 | H |
| (Nagarkar et al., 2022) | Y | N | Y | Y | Y | Y | N | Y | N | Y | N | 7 | M |
| (Nguyen et al., 2021) | Y | Y | Y | Y | Y | N | N | Y | N | N | N | 6 | M |
| (Ning et al., 2020) | Y | N | Y | Y | Y | N | N | Y | N | N | N | 5 | M |
| (Nordin et al., 2022) | Y | N | Y | Y | Y | N | N | Y | N | N | N | 5 | M |
| (NS et al., 2021) | Y | N | Y | Y | Y | N | N | N | N | N | N | 4 | M |
| (Ornell et al., 2020) | Y | N | Y | Y | Y | N | N | Y | N | Y | N | 6 | M |
| (Oteir et al., 2022) | Y | Y | Y | Y | Y | N | N | Y | N | Y | Y | 7 | M |
| (Pan et al., 2022) | Y | Y | Y | Y | Y | N | N | Y | N | Y | N | 7 | M |
| (Pandey et al. 2021) | Y | N | Y | Y | Y | N | Y | N | N | N | N | 5 | M |
| (Paniagua-Ávila et al., 2022 | Y | Y | Y | Y | Y | Y | N | Y | N | Y | N | 8 | H |
| (Pappa et al., 2021) | Y | N | Y | Y | Y | N | N | Y | N | Y | N | 6 | M |
| (Parthasarathy et al., 2021) | Y | N | Y | Y | Y | N | N | Y | N | Y | N | 6 | M |
| (Pelissier et al., 2021) | Y | Y | Y | Y | Y | N | N | Y | N | Y | N | 7 | M |
| (Pérez Herrera et al. 2021) | Y | Y | Y | Y | Y | N | N | Y | N | N | N | 6 | M |
| (Pisanu et al., 2022) | Y | N | Y | Y | Y | N | N | Y | N | N | N | 5 | M |
| (Proserpio et al., 2022) | Y | N | Y | Y | Y | N | N | Y | N | N | N | 5 | M |
| (Rahman et al., 2021) | Y | Y | Y | Y | Y | N | N | Y | N | Y | N | 7 | M |
| (Repon et al., 2021) | Y | Y | Y | Y | Y | N | Y | Y | N | Y | N | 8 | M |
| (Riaz et al., 2021) | Y | N | Y | Y | Y | N | Y | Y | N | Y | N | 7 | M |
| (RilleraMarzo et al., 2021) | Y | Y | Y | Y | Y | N | N | Y | N | Y | N | 7 | M |
| (Rossi et al. 2020) | Y | N | Y | Y | Y | N | N | N | N | N | N | 4 | M |
| (Sabbaghi et al., 2022) | Y | Y | Y | Y | Y | N | N | Y | N | N | N | 6 | M |
| (Salehiniya and Abbaszadeh, 2021) | Y | N | Y | Y | Y | N | N | Y | N | Y | N | 6 | M |
| (Salman et al., 2022) | Y | Y | Y | Y | Y | N | N | Y | N | Y | N | 7 | M |
| (Samir AlKudsi et al., 2022) | Y | N | Y | Y | Y | N | N | Y | N | Y | N | 6 | M |
| (Sertoz et al., 2021) | Y | N | Y | Y | Y | N | N | Y | N | N | N | 5 | M |
| (Setiawati et al., 2021) | Y | N | Y | Y | Y | N | N | Y | N | N | N | 5 | M |
| (Shah et al., 2021a) | Y | N | Y | Y | Y | N | N | Y | N | Y | N | 6 | M |
| (Shah et al., 2021) | Y | N | Y | Y | Y | N | N | Y | N | Y | N | 6 | M |
| (Shah et al., 2020) | Y | N | Y | Y | Y | N | N | Y | N | N | N | 5 | M |
| (Shechter et al., 2022) | Y | N | Y | Y | Y | Y | N | Y | N | Y | Y | 8 | H |
| (Shechter et al., 2020) | Y | N | Y | Y | Y | N | N | N | N | Y | N | 5 | M |
| (Shen et al., 2020) | Y | N | Y | Y | Y | Y | N | Y | N | Y | N | 7 | M |
| (Shrestha, 2020) | Y | N | Y | Y | Y | N | N | N | N | N | N | 4 | M |
| (Siamisang et al., 2022) | Y | N | Y | Y | Y | N | N | Y | N | N | N | 5 | M |
| (Simonetti et al., 2021) | Y | Y | Y | Y | Y | N | N | Y | N | Y | N | 7 | M |
| (Singh et al., 2021) | Y | N | Y | Y | Y | N | N | Y | N | N | N | 5 | M |
| (Sitanggang et al., 2021) | Y | N | Y | Y | Y | N | N | Y | N | Y | N | 6 | M |
| (Smallwood et al., 2021) | Y | N | Y | Y | Y | N | N | Y | N | Y | N | 6 | M |
| (Sobregrau Sangra et al., 2022) | Y | Y | Y | Y | Y | Y | Y | Y | N | N | N | 8 | H |
| (Song et al., 2020) | Y | N | Y | Y | Y | N | N | Y | N | N | N | 5 | M |
| (Srikanth et al., 2022) | Y | N | Y | Y | Y | N | N | N | N | Y | N | 5 | M |
| (Stocchetti et al., 2021) | Y | N | Y | Y | Y | Y | Y | Y | N | Y | N | 8 | H |
| (Su et al. 2021) | Y | N | Y | Y | Y | N | N | Y | N | N | N | 5 | M |
| (Su et al. 2021) | Y | N | Y | Y | Y | N | N | Y | N | N | N | 5 | M |
| (Sun et al. 2020) | Y | N | Y | Y | Y | N | Y | N | N | Y | N | 6 | M |
| (Suryavanshi et al. 2020) | Y | N | Y | Y | Y | N | N | Y | N | N | N | 5 | M |
| (Syamlan et al. 2022) | Y | N | Y | Y | Y | N | Y | Y | Y | N | N | 7 | M |
| (Tan et al., 2020) | Y | N | Y | Y | Y | N | N | Y | N | Y | N | 6 | M |
| (Tan et al. 2021) | Y | N | Y | Y | Y | N | N | Y | Y | Y | N | 7 | M |
| (Tang et al., 2022) | Y | N | Y | Y | Y | N | Y | Y | N | Y | N | 7 | M |
| (Tao et al. 2021) | Y | Y | Y | Y | Y | N | N | Y | N | Y | N | 7 | M |
| (Taşdelen et al., 2022) | Y | N | Y | Y | Y | N | N | Y | N | Y | N | 6 | M |
| (Than et al., 2020) | Y | N | Y | Y | Y | N | N | Y | N | Y | N | 6 | M |
| (Turan et al., 2022) | Y | N | Y | Y | Y | N | N | N | N | Y | N | 5 | M |
| (Urzua et al., 2020) | Y | N | Y | Y | Y | N | N | N | N | N | N | 4 | M |
| (Uyaroğlu et al., 2020) | Y | N | Y | Y | Y | N | N | N | N | N | N | 4 | M |
| (Uz et al., 2022) | Y | Y | Y | Y | Y | N | N | Y | N | Y | N | 7 | M |
| (Vadi et al., 2022) | Y | N | Y | Y | Y | N | N | Y | N | N | N | 5 | M |
| (Valaine et al., 2021) | Y | Y | Y | Y | Y | N | Y | N | N | N | N | 6 | M |
| (van de Venter et al., 2021) | Y | N | Y | Y | Y | Y | N | N | N | N | N | 5 | M |
| (Van Wert et al., 2022) | Y | N | Y | Y | Y | Y | N | Y | N | Y | N | 7 | M |
| (Vancappel et al., 2021) | Y | Y | Y | Y | Y | N | N | Y | N | N | N | 6 | M |
| (Wanigasooriya et al., 2020) | Y | Y | Y | Y | Y | N | Y | Y | N | Y | N | 8 | H |
| (Wayessa et al., 2021) | Y | Y | Y | Y | Y | Y | N | Y | N | Y | N | 8 | H |
| (Weibelzahl et al., 2021) | Y | N | Y | Y | Y | Y | N | Y | N | N | N | 6 | M |
| (Xiong et al., 2020) | Y | N | Y | Y | Y | N | N | N | N | Y | N | 5 | M |
| (Yadeta et al., 2021) | Y | Y | Y | Y | Y | Y | N | Y | N | Y | N | 8 | H |
| (Yang et al., 2022) | Y | Y | Y | Y | Y | Y | N | Y | N | Y | N | 8 | H |
| (Youssfi et al. 2021) | Y | N | Y | Y | Y | N | N | N | N | N | N | 4 | M |
| (Yu et al. 2022) | Y | N | Y | Y | Y | N | Y | Y | N | Y | N | 7 | M |
| (Zarzour et al. 2022) | Y | N | Y | Y | Y | N | N | Y | N | N | N | 5 | M |
| (Zhan et al. 2022) | Y | Y | Y | Y | Y | N | Y | N | N | Y | N | 7 | M |
| (Zhang et al. 2020) | Y | N | Y | Y | Y | N | Y | N | N | N | N | 5 | M |
| (Zhang et al. 2021) | Y | N | Y | Y | Y | N | N | N | N | N | N | 4 | M |
| (Zhang et al. 2020) | Y | Y | Y | Y | Y | N | N | N | N | N | N | 5 | M |
| (Zhang et al. 2022) | Y | N | Y | Y | Y | N | Y | N | N | Y | N | 6 | M |
| (Zheng et al. 2021) | Y | Y | Y | Y | Y | N | Y | Y | N | N | N | 7 | M |
| (Zhu et al. 2020) | Y | N | Y | Y | Y | N | Y | Y | N | Y | N | 7 | M |

Q1:Is the source of the data clear (survey, literature review)?

Q2:Are the inclusion and exclusion criteria for exposure and non-exposure groups (cases and controls) listed or referred to previous publications?

Q3:Is the time period for identifying patients given?

Q4:If it is not the source of the population, are the subjects continuous?

Q5:Did the evaluators’ subjective factors mask other aspects of the study subjects?

Q6:A description of any assessments performed for quality assurance (e.g. testing/retesting of subjective outcome measures)

Q7:Explains the reasons for excluding any patient from the analysis.

Q8:Describes how to evaluate and/or control for confounding measures.

Q9:If possible, explains how missing data is handled in the analysis.

Q10:The response rate of patients and the integrity of data collection are summarized.

Q11:If there is a follow-up, identify the percentage of incomplete data or follow-up results of the expected patients.

**S5 Table: Newcastle-Ottawa observational study scale (NOS)**

| Author/year | Q1 | Q2 | Q3 | Q4 | Q5 | Q6 | Q7 | Q8 | Score | Quality |
| --- | --- | --- | --- | --- | --- | --- | --- | --- | --- | --- |
| (Ali et al., 2020) | 1 | 1 | 1 | 1 | 2 | 1 | 0 | 0 | 7 | H |
| (Martin et al., 2021) | 1 | 1 | 1 | 0 | 2 | 1 | 0 | 0 | 6 | H |
| (McGuinness et al., 2022) | 1 | 1 | 1 | 1 | 2 | 1 | 0 | 0 | 7 | H |

Q1:Representativeness of the exposure cohort.

Q2:Selection of non-exposed cohorts.

Q3:Ascertainment of exposure.

Q4:Demonstration that outcome of interest was not present at start of study.

Q5:Comparability of cohorts on the basis of the design or analysis.

Q6:Assesment of outcome.

Q7:Was following-up long enough for outcomes to occur?

Q8:Adequacy of follow up of cohort.
